# Mapping the Dynamic Interplay of Mental Health and Weight Across Childhood: Data-Driven Explorations Using Causal Discovery

**DOI:** 10.64898/2026.04.16.26350943

**Authors:** Tobias Ellegaard Larsen, María Hernández-Lorca, Claus Thorn Ekstrøm, Rebecca Vinding, Klaus Bønnelykke, Katrine Strandberg-Larsen, Anne Helby Petersen

## Abstract

Childhood weight development, especially overweight and obesity, has been associated with mental health, but their dynamic, causal relationships, and whether these differ by sex, remain unclear. We applied causal discovery to data from the Danish National Birth Cohort (n=67,593) spanning six periods from pregnancy to late adolescence and considering 67 variables related to child and parental weight, mental health, lifestyle, and socio-economic factors. We found no statistically significant difference between the causal graphs for boys and girls (*P*=0.079). The data-driven models found causal influence of childhood weight on subsequent weight status. Mental health pathways were exclusively within or across adjacent periods and centered on early adolescent stress. We examined the interplay between a subset of mental health variables, containing information on externalizing and internalizing problems, and weight, and found no direct causal pathway between the two processes. These findings suggest that observed links between weight and these mental health measures may be attributable to confounding. Our findings demonstrate the value of data-driven causal discovery in large cohort studies and how to test for differences in causal mechanisms across subgroups. Results are available in an interactive application, enabling future research to further explore the interplay between weight and mental health.

## INTRODUCTION

Childhood overweight and obesity have consistently been associated with anxiety, depression, conduct and emotional problems, peer relationship difficulties, reduced quality of life, and low self-esteem in children and adolescents^1–5^. During recent decades, the prevalence of childhood obesity has increased in every world region^6,7^ and the onset has been moving towards younger ages^8,9^. Given that many psychiatric conditions begin in childhood^10^ and commonly co-occur with overweight, examining the temporal interplay between weight and mental health is timely and important^11^.

Prevalence of childhood obesity differs between boys and girls^12^, and the causal interplay between weight and mental health might also operate differently in each sex. In a large birth cohort, the association between higher BMI at age 7 years and greater body dissatisfaction at age 11 years was twice as large in girls than in boys^13^. A multicenter study of children, adolescents and young adults with overweight or obesity and comorbid mental health disorders found that obese males were more likely to present attention-deficit/impulsivity disorder (ADHD) comorbidity, but obese females depression and eating disorders^2^.

Critical biological time windows and life events might be central to the relationship between mental health and weight. Intrauterine factors (e.g., elevated maternal body mass index [BMI]^14,15^ and fetal growth abnormalities^15–17^) and postnatal experiences (e.g., rapid infant weight gain^18,19^, adverse experiences^20,21^, parental mental health problems^22,23^, and breastfeeding^24,25^) are associated with both obesity and mental health outcomes. Prior research highlights social, behavioral^26^, and psychological aspects^13,27–30^ as key contributors to this interplay. Longitudinal evidence also suggests bidirectionality, with early psychological distress and behavioral dysregulation predicting later increases in BMI through pathways such as mental distress^31,32^, ADHD symptoms^33^, sleep disruption^34^, and stress^35^ mechanisms. Understanding these complex pathways, including familial factors that remain underexplored^36^, is critical for identifying intervention targets that could disrupt this cycle and mitigate long-term adverse outcomes. Most studies do not model this complex, dynamic process, but rather focus on single exposure-outcome relationships irrespective of the age span, which limits insight into the full causal system.

Recent advances in causal-discovery machine-learning methods allow for explorations of high-dimensional longitudinal data, facilitating a shift toward system-level mechanistic understanding of complex health phenomena^37–40^. These approaches infer the causal model that best explains observed statistical patterns, leveraging temporal structure in longitudinal data^41–44^. In this study, we conduct fully data-driven explorations of the interplay between weight and various aspects of mental health through childhood and assess whether the discovered mechanisms differ by sex. We include other factors hypothesized to affect this interplay, including health measures, socio-economic factors, childhood adversities, and lifestyle factors, totaling 67 variables. Using high-quality longitudinal data from the Danish National Birth Cohort (DNBC) from pregnancy to age 18 years^45^, we apply a causal discovery algorithm to investigate the complex, time-dynamic causal mechanisms linking weight and mental health across childhood, allowing for sex-specific mechanisms. This approach moves beyond pre-specified causal models (e.g., represented as directed acyclic graphs) and offers the potential to reveal previously unrecognized pathways and mechanistic insights. This allows us to go beyond prior confirmatory research and pose new hypotheses about when and how interventions could most effectively disrupt adverse joint trajectories of weight and mental health.

## METHODS

### Study population

The DNBC is a population-based birth cohort consisting of 96,822 individuals born from 1996 to 2003 (thereof 92,657 liveborn singletons). Pregnant women were enrolled and started providing information during prenatal life^45,46^. The cohort has several follow-ups on both mothers and offspring until age 18 years.

Ethical approvals and data access permissions are described in Web Appendix S1.

*Inclusion criteria*: Liveborn singletons, residence in Denmark and alive until age 18 years and 3 months and participation in the first (prenatal) DNBC interview.

*Exclusion criteria*: Individuals with congenital anomalies diagnosed before 1 year of age, conditions affecting the central nervous system diagnosed before 7 years of age, a global developmental delay diagnosis, endocrine and metabolic diseases (see supplement for details and ICD-10 codes), having no registered father, experiencing death of a parent before age 18 years and 3 months, born before gestational week 28 and/or born with birth weight <1500 g.

Figure S1 summarizes the exclusion process, which results in a study population of *n =* 75103 individuals, hereof *n_girls_ =* 36458 and *n_boys_* = 38645.

We base the analysis on a randomly selected sample of 90% of the study population meeting the criteria above, allowing the remaining 10% to be used for future research without imposing risks of overfitting.

### Measurements

Table 1 summarizes all variables included (see Table S1 for details). Each variable was assigned to a temporal period (hereafter *tiers)*: i) *Pregnancy* (one year before birth to 16 weeks of gestation); ii) *At birth*; iii) *Infancy* (6-18 months); iv) *Childhood* (7 years); v) *Early adolescence* (11 years); and vi) *Late adolescence* (18 years). All variables were further classified into categories i) *Weight* (child and parental weight); ii) *Mental health* (child’s mental health); iii) *Lifestyle risk factors* (selected behavioral factors that may be targeted in interventions); or iv) *Other* (none of the above, e.g., possible confounders).

**Table 1:**
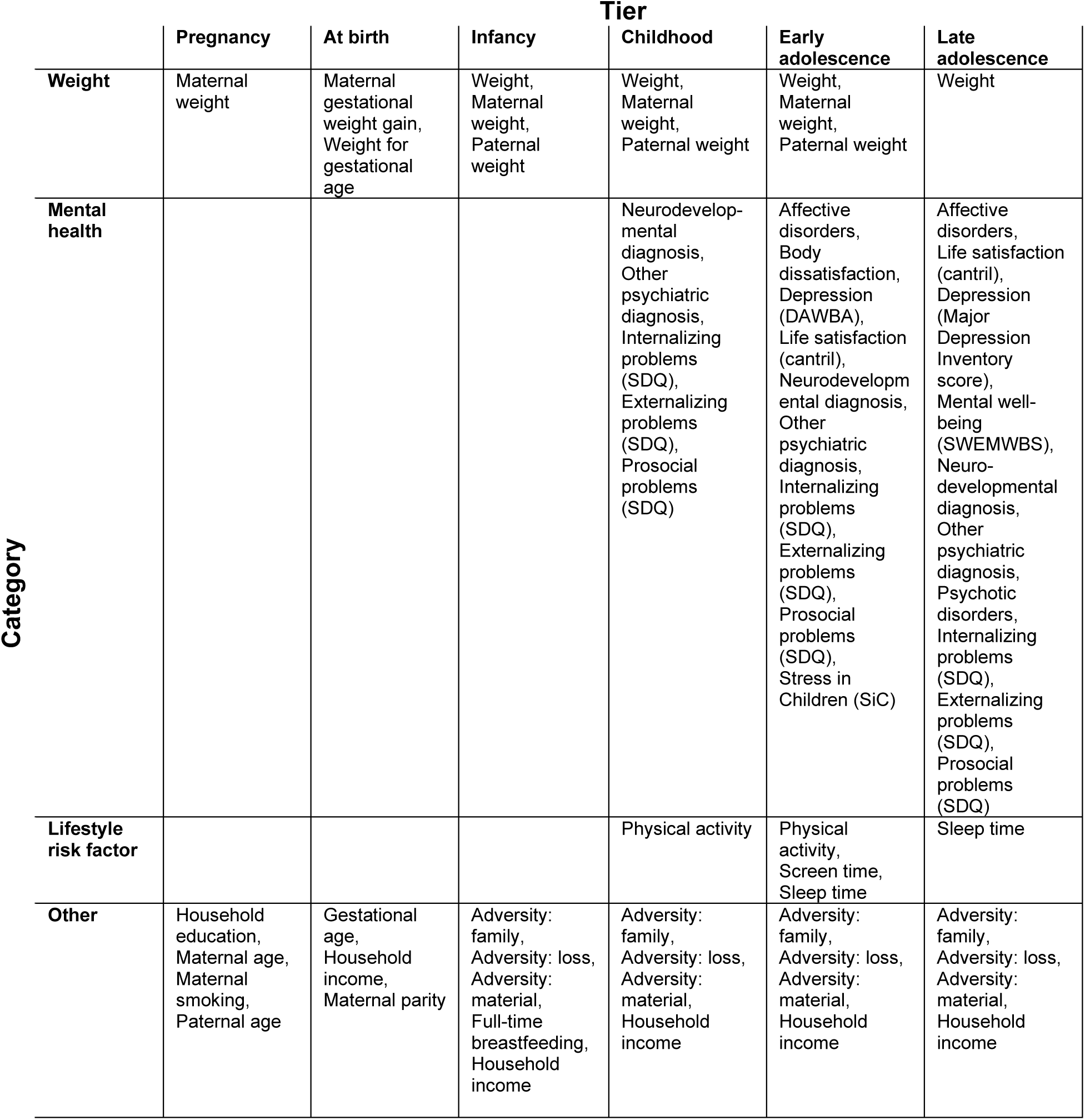
Overview of variables included, organized according to category and tier. Variables refer to the offspring unless otherwise stated. Abbreviations: SWEMWBS - Short Warwick-Edinburgh Well-Being Score; SDQ – Strengths and Difficulties Questionnaire, SiC – Stress in Children questionnaire; DAWBA - Development and Well-Being Assessment

### Statistical analysis

#### Descriptive statistics

All variables are categorical. We reported marginal distributions as relative frequencies stratified by sex, along with the amount of missing information, based on three nested populations:

1. **Full study population**: DNBC participants meeting the inclusion/exclusion criteria from Section 2.1.
2. **Complete participants**: Members of the full study population that participated in all data collections.
3. **Analysis sample**: Complete participants and (weighted) clones of complete participants, see details below.

This allowed for assessing possible selection issues.

#### Causal discovery

We used the temporal greedy equivalence search (TGES) causal discovery algorithm to find the family of causal mechanistic models that best explains the data, assuming there are no unobserved confounders^44^. TGES searches for such a model family while enforcing a tiered ordering: potential causes MUST temporally precede their effects. TGES considers several directed acyclic graph (DAG) candidates, computes a statistical score for each, and identifies the family of DAGs that best fits the data. This estimated family is provided as a *temporal maximally oriented partially directed acyclic graph* (TMPDAG). A TMPDAG represents the set of DAGs that are consistent with the observed data and temporal ordering. A TMPDAG is interpreted similarly to a DAG, except that some edges are undirected, reflecting that it is not possible to determine the direction of the causal relationship from observational data and temporal ordering alone.

Given a sufficiently large sample size (possibly infinite), TGES will identify the correct and maximally informative TMPDAG^44^. This means that it makes no false causal claims (i.e., it will not infer that X causes Y if it is not truly the case), and that no additional causal information can be inferred from the data and temporal structure without making additional assumptions (i.e., if the there is an undirected edge between X and Y then the direction of this relationship cannot be determined without making additional assumptions). However, when using empirical data, and hence finite sample sizes, the algorithm may make some errors due to statistical uncertainty.

#### Implementation details

TGES requires the choice of a statistical score criterion, which provides a score for each considered DAG. Since all variables are categorical, we used a discrete Bayesian Information Criterion (BIC) score, which depends on the multinomial maximum likelihood for the DAG, as well as a penalty term for the number of parameters of this model. The size of the penalty term affects how many edges will be in the final estimated TMPDAG. We tuned TGES to find a penalty term that resulted in a TMPDAG with exactly 300 edges. This target number was selected based on simulations and preliminary analysis results that made the level of graph connectedness plausible and informative, focusing on meaningful connections between weight and mental health (details in Web Appendix S2).

The data contains two types of missing information, each addressed with a different strategy:

1. Partial missing information within a data collection, e.g., omitted items or erroneous information.
2. Missing information due to skipping one or more data collections (including dropout).

Partial missing information was handled using single imputation with random forests (details in Web Appendix S3). To account for non-random selection mechanisms in skipping data collections, we used data cloning (details in Web Appendix S4). In a sensitivity analysis, we also considered only the complete participants without cloning.

To assess the stability of the entire analysis procedure (imputation, data cloning and TGES) we conducted 100 bootstrap repetitions using random sampling with replacement (details in Web Appendix S6).

The entire analysis procedure was conducted separately for boys and girls. We used the *Structural Hamming Distance* (SHD) as a measure of overall differences between the TMPDAGs for boys and girls. The SHD counts the number of graph modifications (edge additions, removals, or orientation reversals) that are needed to transform one graph into the other^47^. To test whether the TMPDAGs for boys and girls were statistically significantly different, we used a *permutation test* (details in Web Appendix S7).

All computations were conducted in R version 4.4^48^. For the TGES computations, we used the *causalDisco* package^49^.

#### Reporting

We reported the full TMPDAGs for boys and girls, as well as three derived subgraphs (also stratified by sex) to enhance readability:

1. Weight development subgraph: includes only weight variables (both offspring and parental).
2. Mental health development subgraph: includes only mental health variables.
3. Weight, internalizing/externalizing problems and lifestyle risk factor subgraph: includes offspring weight variables, internalizing problems, externalizing problems, and lifestyle risk factors.

In these subgraphs, an edge is drawn between two variables whenever a (partially) directed path exists between them in the full TMPDAG, see details in Web Appendix S5. Thus, the subgraphs display (possible) flow of causal information among variables of interest, but they do not represent, for example, whether causal pathways are confounded. We only included edges for pathways present in a minimum 80% of the bootstrap repetitions. The first two subgraphs display the interplay within the weight and well-being processes separately. The purpose of including the third subgraph is to showcase how a targeted subgraph analysis can help inform intervention design and etiological considerations.

To allow for further exploration of the results, we developed an interactive web application, which displays the full TMPDAGs stratified by sex. It allows the user to highlight edges shared between sexes, highlight all directed paths between two variables, create subgraphs across variables of interest, or compute the percentage of bootstrap repetitions in which there exists a directed path between two variables.

The web application was developed in R using the *shiny* package^50^ and can be found online at https://shiny.sund.ku.dk/InteractiveGraph/. TMPDAGs and subgraphs were plotted in R using the *causalDisco* package^49^.

## RESULTS

### Descriptive statistics

Table 2 shows the number of DNBC participants included in each tier after applying the inclusion criteria. The number of participants excluded according to each criterion is detailed in Figure S1.

**Table 2:**
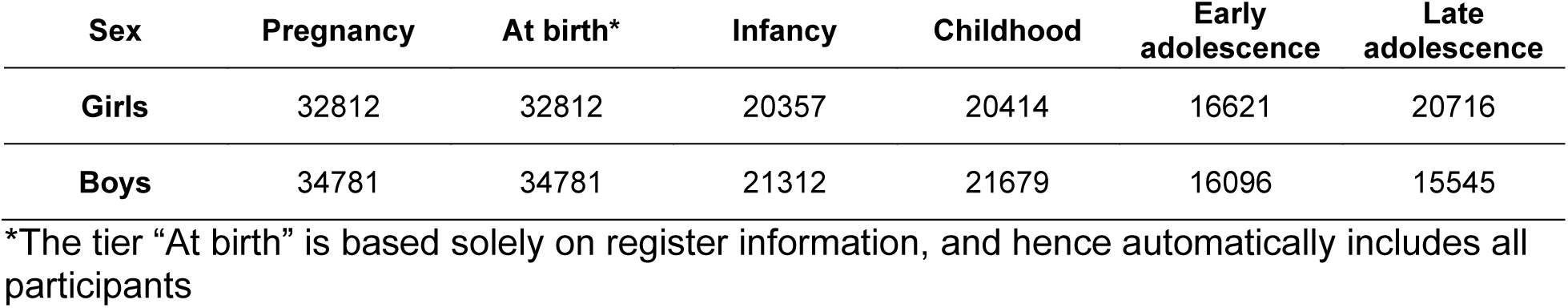
Number of DNBC participants in each tier.

Table S2 presents the distributions of all included variables across three study populations: the full study population, the complete participants, and the analysis sample (with cloning to account for selection), each stratified by sex.

In Table S3 we report the number of unique edges across repetitions and how many of these were present in the TMPDAGs derived from the main analysis.

### Causal mechanisms

Figure 1 shows the TMPDAGs, each with 300 edges, obtained from the temporal causal discovery procedure for boys and girls, respectively. For boys, only 3 edges in the TMPDAG cannot be oriented, while there were 10 unoriented edges among girls. There are 142 shared edges with the same orientation for boys and girls, and 20 additional edges are shared for boys and girls but with different orientations. Using our developed web application the reader can interactively explore the causal models further: https://shiny.sund.ku.dk/InteractiveGraph/.

**Figure 1:**
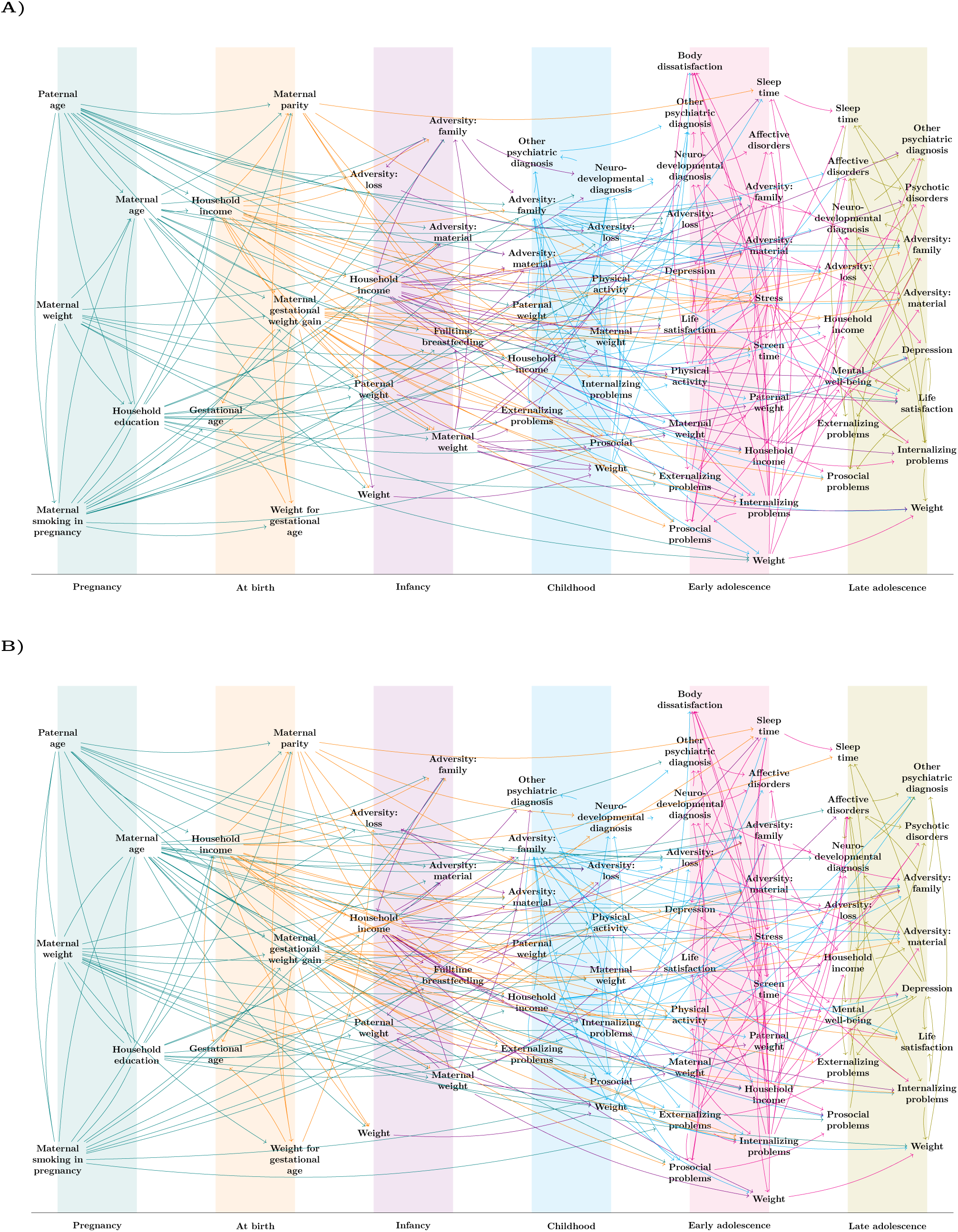
TMPDAGs for boys (A) and girls (B). Edges are colored according to the tier from which they originate. Undirected edges means that the causal discovery algorithm cannot determine their orientation from data and temporal background information alone.

Comparing the TMPDAGs for boys and girls resulted in SHD = 296. The test for no difference in graph structure between boys and girls yielded a *P*-value of *P* = 0.079, and we therefore do not have evidence of any statistically significant difference in the graph structures, testing at a 5% level.

For boys, all 300 edges were observed in at least one bootstrap sample, whereas for girls, two edges were absent across all bootstrap replications. Moreover, all edges, for both boys and girls, that appeared in at least 95% of the bootstrap samples were also present in the TMPDAGs derived from the main analysis.

#### Weight development

Figure 2 presents weight development subgraphs for boys and girls. We identified 22 shared oriented edges, while 5 edges were unique to girls and 2 edges unique to boys. Thus overall, most causal links between weight variables were shared for boys and girls. In Figure S2 and S3 we present the weight development subgraphs, had the threshold for pathways included been set to 50% or 95% instead. The number of pathways is reported in Table S4.

**Figure 2:**
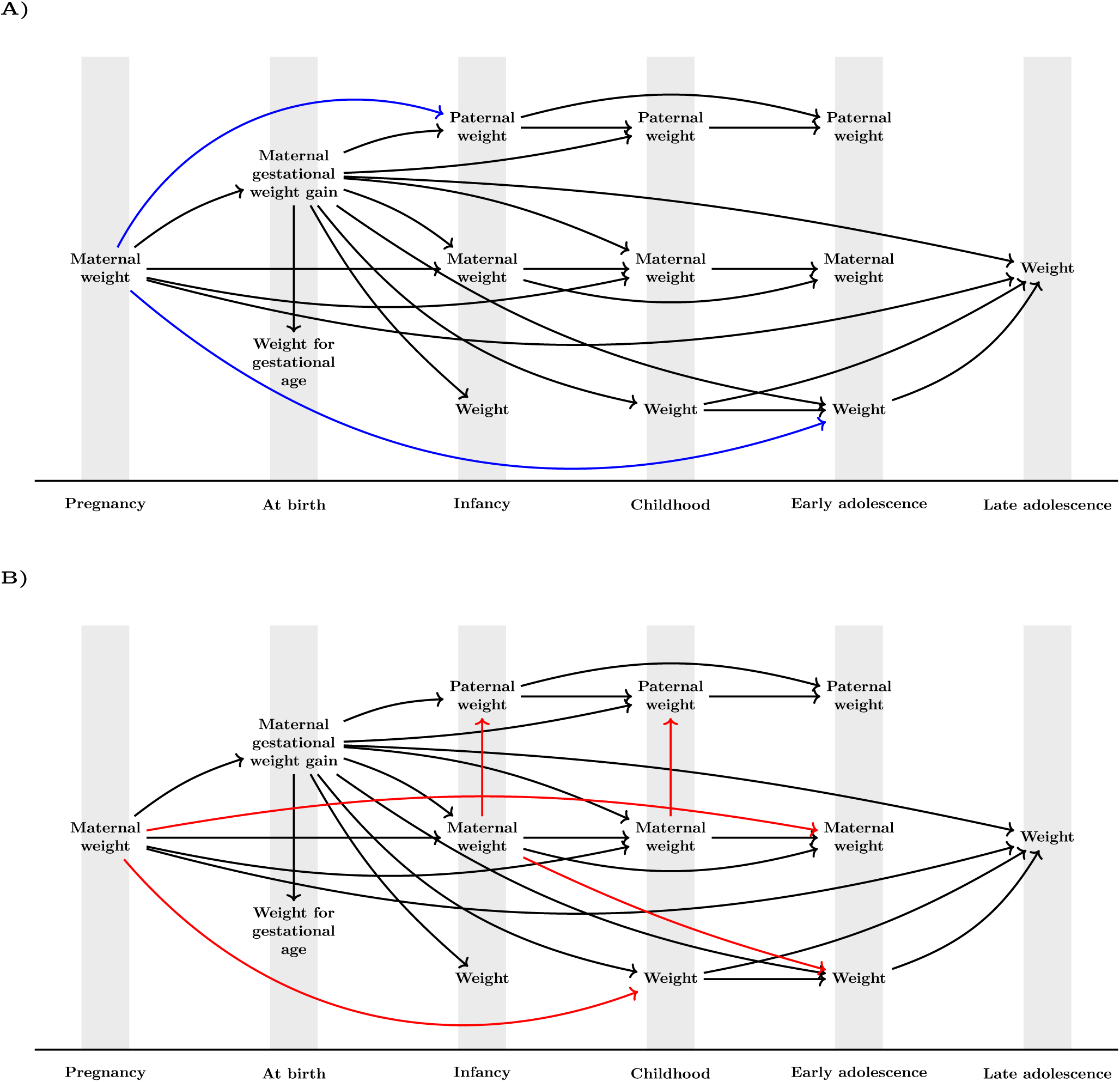
Weight development processes for boys (A) and girls (B). Black edges are shared by boys and girls; red edges are unique to girls and blue are unique to boys. Directed edges represent directed causal paths that are not exclusively mediated via any variables shown in the graph. Undirected edges represent undirected or partially directed causal paths that are not exclusively mediated via any variables shown in the graph. No undirected edges were found.

We observed that offspring weight measures are causally connected from childhood through early adolescence to late adolescence. Interestingly, we also found a lagged effect, childhood weight → late adolescence weight, suggesting a direct causal pathway that is not fully mediated through early adolescence weight. We found no direct causal pathways from either weight for gestational age at birth or weight at infancy to any temporally succeeding weight.

Maternal weight acted as a cause of paternal weight and offspring weight, although the specific patterns differed for boys and girls. Girls appeared more influenced by maternal weight, with girls’ weight in both childhood and early adolescence influenced by, respectively, maternal weight in pregnancy and infancy.

Finally, maternal gestational weight gain and maternal weight in pregnancy were causes of many subsequent weight values for mothers, fathers and offspring, although the specific pathways differed slightly for boys and girls. We especially found maternal gestational weight gain to have causal influence on offspring weight in all later tiers.

#### Mental health development

Figure 3 shows mental health development processes for boys and girls. The graph for boys contains 34 edges representing causal pathways between mental health variables, whereas the graph for girls contains 26. Of these, 17 were identical for boys and girls. Overall, we found a large overlap in the processes for boys and girls, though with more sex-specific differences than for weight development. In Figure S4 and S5 we present the mental health development subgraphs, had the threshold for pathways included been set to 50% or 95% instead. The number of pathways for these are reported in Table S4.

**Figure 3:**
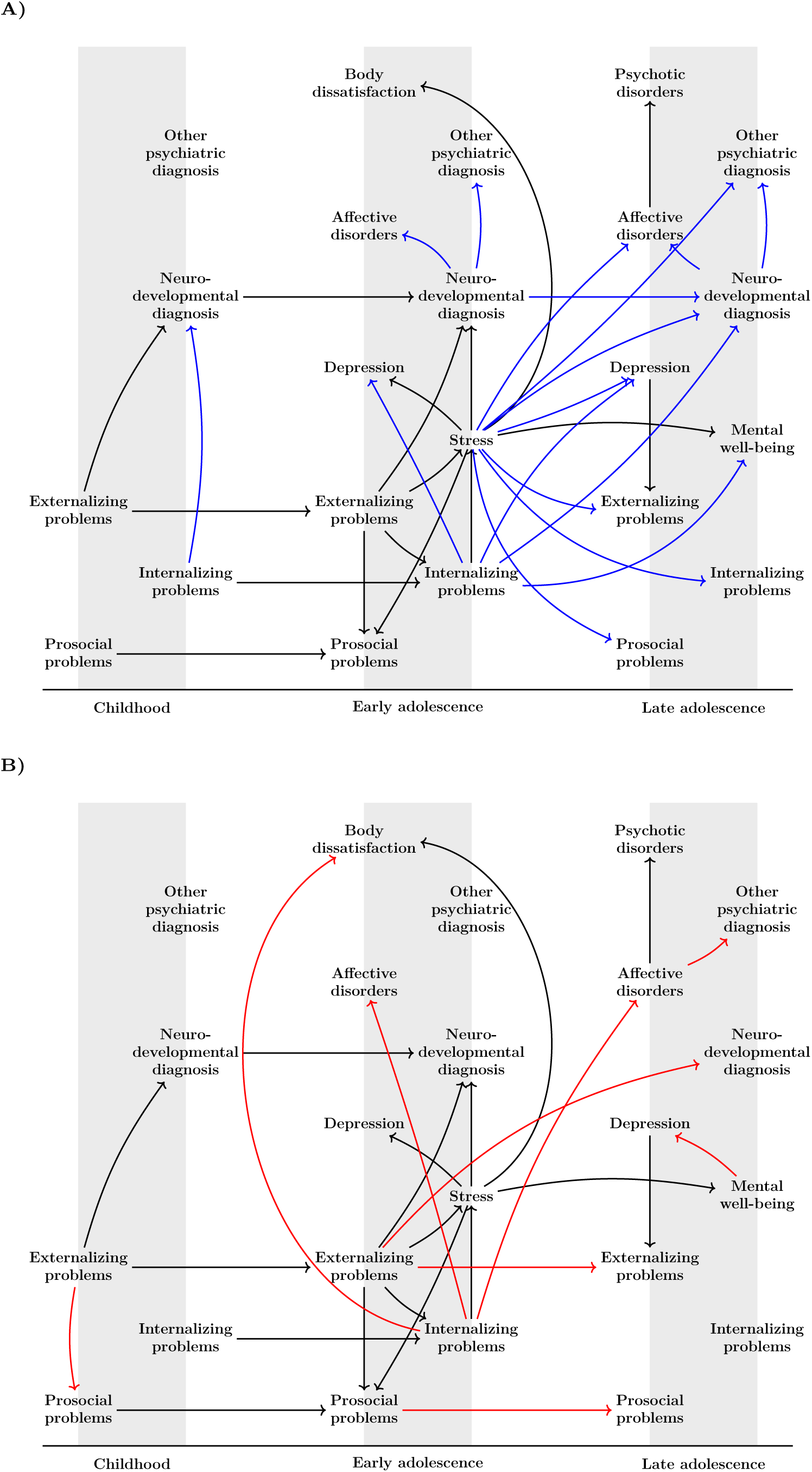
Mental health development processes for boys (A) and girls (B). Black edges are shared by boys and girls; red edges are unique to girls and blue are unique to boys. Directed edges represent directed causal paths that are not exclusively mediated via any variables shown in the graph. No undirected edges were found.

We found no direct causal paths from childhood to late adolescence that were not mediated through early adolescence. For especially boys, stress and internalizing problems in early adolescence appeared to be key nodes through which a vast number of causal pathways went to or originated from.

#### Interplay between lifestyle, weight and internalizing/externalizing problems development

An example of a focused analysis of two of the mental health measures, internalizing and externalizing problems, and their longitudinal interplay with weight development and selected lifestyle risk factors is presented in Figure 4. In Figure S6 and S7 we present the subgraph had the variables been the same but the threshold for pathways included been set to 50% or 95% instead.

**Figure 4:**
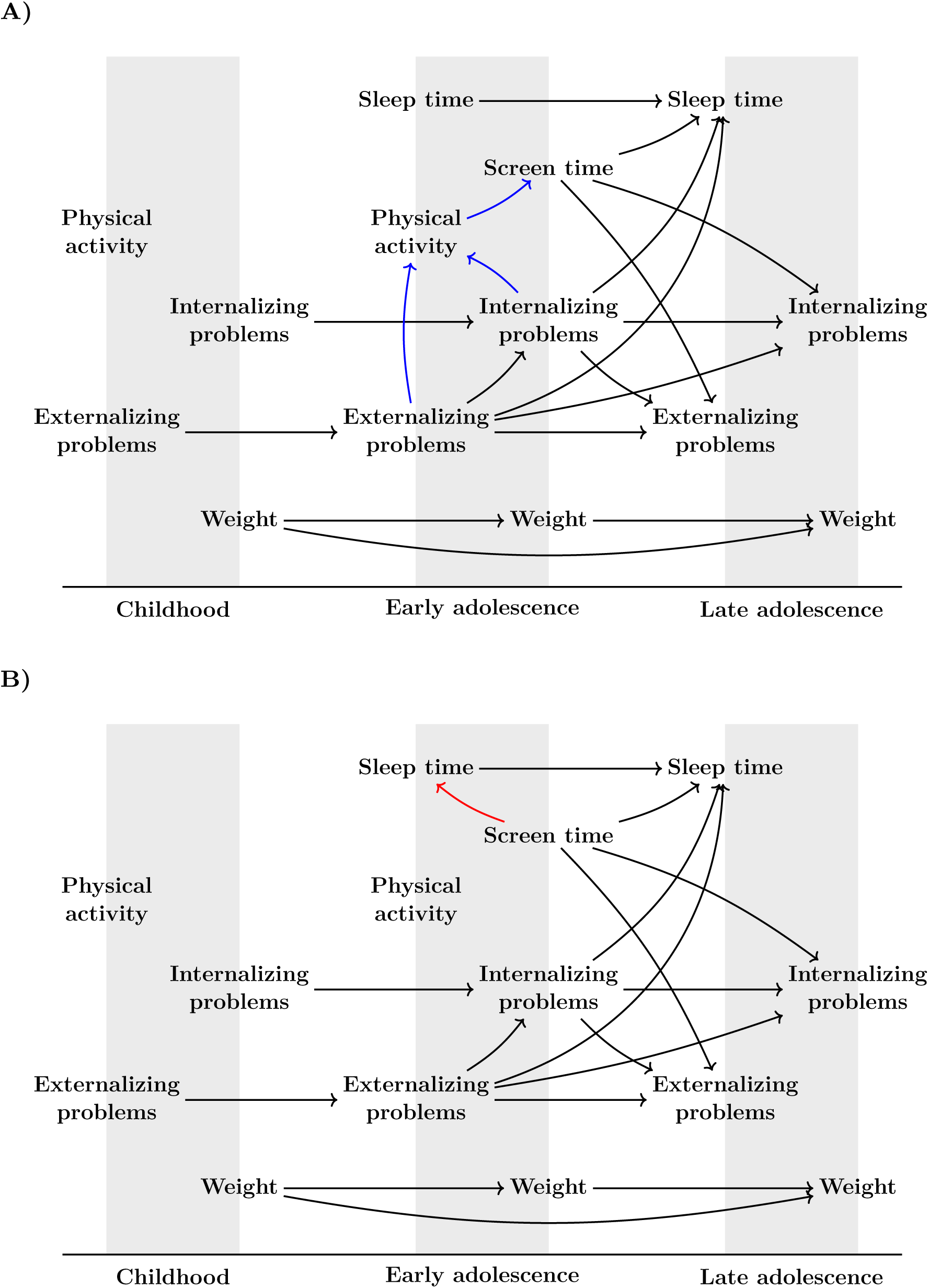
Interplay between weight development, internalizing/externalizing problems and selected lifestyle risk factors for boys (A) and girls (B). Black edges are shared by boys and girls; red edges are unique to girls and blue are unique to boys. Directed edges represent directed causal paths that are not exclusively mediated via any variables shown in the graph. No undirected edges were found. We omit variables from “At birth” and “Infancy” as there were no paths between these and any other node in the graph.

We found identical weight development and internalizing/externalizing problem processes for boys and girls, with no direct linkage between weight and either internalizing/externalizing problems or lifestyle risk factors. This graph does not include shared common causes that can account for any association between weight and mental health. Screen time was the only lifestyle risk factor with a direct causal path to internalizing/externalizing problems. Any differences between sexes are found in relation to lifestyle factors.

#### Sensitivity analysis using complete participants

We compared the TMPDAG obtained from the analysis sample with TMPDAGs obtained from the complete participant sample (details in Web Appendix S6). For boys, this resulted in 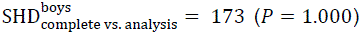 and for girls, we found 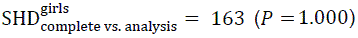. In doing this, we found a larger SHD between the graphs on the analysis sample and the bootstrap samples than we did between the analysis sample and complete participants and hence, we found no statistically significant differences in the estimated TMPDAGs between data cloning and complete participants.

## DISCUSSION

Using temporal causal discovery, we analyzed sex-specific causal pathways connecting weight, mental health, and lifestyle factors from pregnancy through childhood. We found similar weight processes for boys and girls, some overlap in mental health processes, and no statistically significant difference between their causal mechanisms. Screen time in early adolescence emerged as a lifestyle risk factor influencing externalizing/internalizing problems in late adolescence. For both sexes, we observed no direct causal paths from externalizing or internalizing problems to weight, or vice versa.

In this study, we explicitly tested the traditional implicit assumption that the underlying causal mechanisms are identical across sexes. Although we did not find statistically significant evidence of this difference, we chose to present sex-stratified results in line with our *a priori* hypothesis. Although our primary hypothesis of overall sex differences in the full causal graphs was rejected at a 5% significance level (*P* = 0.079), it does not rule out differences in specific subprocesses.

The time-lagged pathway from childhood weight to late-adolescent weight, not fully mediated by early-adolescent weight, is consistent with the idea of critical or sensitive periods in weight development^51^. The absence of any direct pathways from weight for gestational age and infancy weight suggests that neither the relative size at birth nor weight at infancy influence later weight causally; rather, relationships between these factors are explained by common causes such as maternal weight.

We found an influence of maternal weight, but not paternal weight, on offspring weight. Consistent with evidence that maternal gestational weight gain affects weight from birth through childhood^52^, our data-driven study found early-life effects and additionally identified influences extending into early and late adolescence. These findings suggest maternal weight during and after pregnancy as potential leverage points for population-level weight modification.

Causal pathways among mental health variables occurred only within or between adjacent tiers, indicating a more rapid and potentially more malleable dynamic than observed for weight development. This suggests interventions could modify mental health outcomes within brief follow-up periods, rather than requiring early life interventions. Our findings identified stress, primarily for boys, as a key point and the breadth of outgoing edges suggests that reducing stress could potentially produce wide-reaching effects across multiple mental health domains. The importance of stress is consistent with existing longitudinal evidence linking adolescent, especially school-related, stress to later mental health difficulties^53–55^. The stress and internalizing problems are connected phenomena with slight overlap in items used to measure these constructs, and the sex-specific differences are likely to reflect sex-specific differences in reporting of symptoms.

When examining the interplay between lifestyle, weight and internalizing/externalizing problems, we found that the only upstream connection of externalizing and internalizing problems in late adolescence to a lifestyle risk factor was screen time. Interventional and observational evidence suggests that not just screen time^56^, but also other behaviors do relate to weight, although real-world effects are often weak, inconsistent, or hard to sustain^57–60^.

Our study found no direct causal paths from externalizing or internalizing problems to weight, and vice versa, indicating that any co-occurrence of these mental health problems and weight can be explained by confounding. This could be attributed to genetic confounding.

As shown in Table S3, repeating our analysis on the bootstrap samples resulted in 1269 (1294) unique edges found for girls (boys), of which 125 (106) appeared in ≥75 % of the sample and 51 (39) appeared in every sample. This indicates few strong and many possible mechanisms.

A recent study has also employed causal discovery methods to identify potential modifiable risk factors causing obesity in children based on the IDEFICS/I. Family cohort^61^. In this study, sex was included as a variable in the models, instead of used for stratification. The analyses included physical activity, sleep, diet, well-being and audio-visual media consumption as risk factors, which were found to have stable indirect effects on weight.

Several inherent limitations of causal discovery with the TGES algorithm should be noted. First, the estimated causal graphs (TMPDAGs) represent DAG families, and a few edge orientations remain unknown, even with temporal constraints. These should be interpreted as ambiguous causal directions, not as bidirectional feedback mechanisms. This poses challenges for causal effect estimation, as a DAG family can contain several DAGs, each of which gives rise to a different adjustment set for a specific causal effect. This is, however, resolvable using the *intervention with DAG absent* framework, which allows for bounding causal effects estimated from DAG families^62^. Second, TGES relies on the assumption of no unmeasured confounding. When this assumption is not fulfilled, the algorithm may find spurious edges and make orientation errors^63^. Alternative algorithms, such as *fast causal inference*^63^, relax this assumption but come at the cost of a resulting causal model that is markedly more difficult to interpret, and computationally very heavy to estimate. Finally, although causal discovery is exploratory, it should also be stressed that it is indeed data-driven: we can only identify links between the variables included, and results should be interpreted with this in mind. For example, we cannot discuss the role of diet in this study, as this variable was not available for the discovery algorithm.

We used only the temporal ordering of the variables for the causal discovery, not other background knowledge. Although we *a priori* deemed some causal links necessarily present, e.g., weight from one tier to the next, we deliberately decided not to impose such expectations in the causal discovery algorithm because the full lag-structure of weight processes was unknown. In fact, a key part of our research question was to determine this lag-structure. Keeping the procedure completely data-driven lets us determine the most stable and strong weight links, gain deeper insights into the developmental process, and allows us to assess the validity of critical period theories, which would not have been possible if we had imposed certain weight links *a priori*.

To our knowledge, this study is the first to systematically use data-driven methods to compare causal mechanisms across subgroups such as boys and girls. The methodology and testing strategies used here can be applied to other grouping variables and hence help generate evidence regarding the appropriateness of grouped causal analysis. Differences found between subgroups would motivate subgroup specific identification of causal effects, selection of adjustment sets, and intervention designs.

While DNBC offers large-scale high-quality longitudinal data, generalizability to more diverse populations would require external validation. Future work should test the data-driven hypotheses generated in external cohorts as well as explore whether incorporating additional environmental or biological measures reveal modifiable upstream causes that are not captured in the present study. Exploring the role of genetics would provide an important additional perspective.

A central novelty and contribution of this study is the application of temporal causal discovery to a large birth cohort with repeated measures from pregnancy through late adolescence, thus demonstrating the feasibility of temporally constrained causal discovery in a large birth cohort. By searching through a high-dimensional, but temporally restricted, space of causal models, we could recover mechanistic structures across multiple domains (weight, mental health, and lifestyle) in one analysis. This is particularly valuable in cohort studies like the DNBC, where complex, interdependent processes unfold over long periods. Such an analysis needs to include many variables to justify the assumption of no unobserved confounding. As shown in Figure 1, the resulting graph is too complex to interpret directly. Therefore, we provided several methodological innovations to address this general issue in causal discovery applications: First, we developed a novel subgraph construction procedure, which allows the user to zoom in on selected variables and their causal pathways, while retaining causal information flow as displayed in the full TMPDAG. Second, we developed an interactive web application that can be used to query the estimated TMPDAGs or construct new subgraphs of interest. We believe that the results from this study can generate a multitude of new hypotheses beyond those already explored here.

## Acknowledgement

Tobias Ellegaard Larsen acknowledges funding from the Danish National Research Foundation, Carlsberg Foundation, Lundbeck Foundation, Novo Nordisk Foundation and Villum Foundation in support of the Pioneer Centre for SMARTbiomed.

## Data availability statement

All datasets generated during the current study and/or analyzed during the current study are not publicly available.

## ABBREVIATIONS

ADHD: Attention Deficit/Hyperactivity Disorder
BIC: Bayesian Information Criterion
BMI: Body Mass Index
DAG: Directed Acyclic Graph
DAWBA: Development and Well-Being Assessment
DNBC: Danish National Birth Cohort
ICD-10: International Statistical Classification of Diseases and Related Health Problems 10th Revision
MDI: Major Depression Inventory
TMPDAG: Temporal Maximally oriented Partially Directed Acyclic Graph
SDQ: Strengths and Difficulties Questionnaire
SHD: Structural Hamming Distance
SiC: Stress in Children questionnaire
SWEMWBS: Short Warwick-Edinburgh Well-Being Score
TGES: Temporal Greedy Equivalence Search

