## Supplementary Material for "Mapping the Dynamic Interplay of Mental Health and Weight Across Childhood: Data-Driven Explorations Using Causal Discovery"

(†) Shared last authorship.

1. Pioneer Centre for SMARTbiomed, Section of Biostatistics, Department of Public Health, Faculty of Health and Medical Sciences, University of Copenhagen, Øster Farimagsgade 5, 1353 Copenhagen K, Denmark

2. Section of Biostatistics, Department of Public Health, Faculty of Health and Medical Sciences, University of Copenhagen, Øster Farimagsgade 5, 1353 Copenhagen K, Denmark

3. Center for Neuropsychiatric Schizophrenia Research (CNSR), Mental Health Center Glostrup, Copenhagen University Hospital – Mental Health Services CPH, Copenhagen, Denmark

4. Copenhagen Prospective Studies on Asthma in Childhood (COPSAC), Copenhagen University Hospital - Herlev and Gentofte, Copenhagen, Denmark.

5. Department of Pediatrics and Adolescent Medicine, Copenhagen University Hospital - Herlev and Gentofte, Herlev, Denmark

6. Department of Clinical Medicine, Faculty of Health and Medical Sciences, University of Copenhagen, Copenhagen, Denmark

7. Section of Epidemiology, Department of Public Health, Faculty of Health and Medical Sciences, University of Copenhagen, Øster Farimagsgade 5, 1353 Copenhagen K, Denmark

8. Copenhagen Health Complexity Center, Department of Public Health, Faculty of Health and Medical Sciences, University of Copenhagen, Øster Farimagsgade 5, 1353 Copenhagen K, Denmark

**(*) Corresponding author**
Tobias Ellegaard Larsen, Section of Biostatistics, Department of Public Health, Faculty of Health and Medical Sciences, University of Copenhagen, Øster Farimagsgade 5, 1353 Copenhagen K, Denmark.

#### List of included materials

Web Appendix S1: Ethics approval and data access

Web Appendix S2: Further details regarding choosing number of edges

Web Appendix S3: Partial missing information handling

Web Appendix S4: Data cloning for addressing selection bias due to individuals skipping data collections or dropping out

Web Appendix S5: Subgraphs for visualization

Web Appendix S6: Bootstrap repetitions

Web Appendix S7: Permutation test

Exclusion criteria

Table S1: Categorization and description of measurements included in the data analysis

Table S2: Distributions of all included variables and levels in the study population

Table S3: Number of unique edges across bootstrap repetitions.

Table S4: Number of pathways between all weight variables and all well-being variables.

Table S5: Data collection skipping patterns observed in the study population.

Figure S2: Data flowchart

Figure S2: Weight development processes for boys and girls with edges for pathways present in over 50% of the bootstrap repetitions.

Figure S3: Weight development processes for boys and girls with edges for pathways present in minimum 95% of the bootstrap repetitions.

Figure S4: Mental health development processes for boys and girls with edges for pathways present in over 50% of the bootstrap repetitions.

Figure S5: Mental health development processes for boys and girls with edges for pathways present in minimum 95% of the bootstrap repetitions.

Figure S6: Interplay between weight development, internalizing/externalizing problems and selected life-style risk factors for boys and girls with edges for pathways present in over 50% of the bootstrap repetitions.

Figure S7: Interplay between weight development, internalizing/externalizing problems and selected life-style risk factors for boys and girls with edges for pathways present in minimum 95% of the bootstrap repetitions.

Figure S8: Subgraph example.

References for supplementary material

#### Web Appendix S1: Ethics approval and data access

The DNBC cohort is approved by the Danish Data Protection Agency under the general approval (fællesfortegnelse) granted to Statens Serum Institut, reference number 18/04608 and by the Committee on Health Research Ethics (case number (KF) 01-471/94). Women that enrolled themselves and their pregnancy in the DNBC provided informed consent, and individuals born into the cohort were notified about their participation, rights and the option to opt out upon turning 18.

Data approvals for the analyses conducted in this study, which involved linking DNBC data with register data accessed on Statistics Denmark's server, were obtained from the DNBC managerial team (2024-20), Statistics Denmark, the Danish Health Data Authority, and registered under the Danish Data Protection Agency's general approval for the Faculty of Health and Medical Sciences at the University of Copenhagen (514-0400/19-3000).

#### Web Appendix S2: Further details regarding choosing number of edges

To choose the number of edges to include in the TMPDAGs, we simulated random TMPDAGs with similar number and tier allocation of variables as in our data, but different numbers of edges (ranging from no edges to fully connected). We then assessed what number of edges resulted in a graph that appeared similar in sparsity to DAGs from published literature, but also not overly complex. The TMPDAGs were displayed in a preliminary version of the interactive app used to visualize the main results. Based on the simulations, we chose 200 edges as a reasonable number. However, this number was subsequently adjusted to 300 after inspecting results from preliminary causal discovery applications on a crude version of the complete case dataset. Here, we observed that 200 edges resulted in only very few edges connecting mental health and weight variables. Based on such a graph, it would be difficult to determine whether a lack of connection reflected that we had imposed too few edges in the graph (i.e. a design artefact) or a real finding. Therefore, we decided to increase the number of edges to 300, thus making it more plausible that if we again observed few edges between mental health and weight, it is less likely to be a design artefact.

#### Web Appendix S3: Partial missing information handling

We imputed partial missing information using single imputation. We used random forest models for the imputation models with 1000 trees. The imputations were conducted in R using the missForest package[1].

We used single imputation rather than multiple imputations since there do not currently exist methods to combine (temporal) GES with multiple imputations. Using single imputation will generally result in variance deflation, as the procedure does not account for additional variability in the data due to imputing missing values[2]. It is not known how this may affect causal discovery performance and should hence be considered a methodological limitation of the procedure.

#### Web Appendix S4: Data cloning for addressing selection bias due to individuals skipping data collections or dropping out

For each possible data collection skipping pattern (e.g., skipping data collections from tiers 1, 2 and 4), we considered the individuals with this skipping pattern as well as *complete participants* who participated in all data collections, and used a random forest model to compute probabilities of this skipping pattern. We used all variables from all other non-skipped tiers, as well as register-based information from the skipped tiers. We then used the inverse probabilities minus 1 to up-sample clones among the complete participants. This resulted in a dataset (the analytical sample) consisting of complete participants and non-randomly selected clones thereof.

More specifically, we proceeded as follows: Divide the dataset into 16 subsets, one for each distinct data collection skipping pattern, see Table S5. Note that tier "at birth" is register-based and hence cannot be skipped, and that we required all participants to have information for the "pregnancy" tier as a study inclusion criterion. Hence these two tiers are not included in any skipping patterns.

We then carried out the partial missingness imputation procedure outlined in Web Appendix S2 separately for each pattern of missingness due to individuals skipping data collections or dropping out, as we hypothesize there may be different partial missing mechanisms for the different skipping pattern subpopulations. For example, for individuals with no skipped data collections, we fitted imputation models only on that subpopulation.

After imputing partial missing information, we used the inverse probability minus 1 weights to upsample the dataset to account for selection bias resulting from individuals that skip one or more data collections: For each of the patterns $i \in 1, \ldots, 15$ from Table S5, we proceeded as follows (separately for boys and girls):

1. We construct a dataset D by combining the complete participant dataset $D_{\text{full}}$ with the skip pattern dataset $D_i$: $D = D_{\text{complete}} \cup D_i$.
2. For each individual in *D,* we use all information available from data collections not skipped in skip pattern i, as well as register-based information from skip pattern *i* (if available) to compute the probability of belonging to dataset $D_i$ (i.e., not being a complete participant). To compute the probabilities, we use random forest models with 10,000 trees in R using the ranger package[3].
3. Let $n_i$ denote the number of individuals with skip pattern *i*. We sample $n_i$ individuals with replacement from $D_{\text{complete}}$ by using the inverses of the probabilities computed in step 2 minus 1 as weights for the sampling. Hence, we are more likely to sample complete participants that resemble those with skip pattern *i* than those who do not. We add these cloned individuals to the final analysis sample.

After repeating these steps for each skipping pattern, we also added the complete participants to the final analysis sample. This sample has $n = n_{\text{complete}} + n_1 + n_2 + \cdots + n_{15} = 34781$ observations for boys.

Note that this data cloning strategy will result in variance deflation, as cloned individuals can appear several times in the data, and hence the analysis sample has less heterogeneity than would be expected if we had had access to full information from all participants. However, we do not have methods for addressing this issue in causal discovery applications, and hence it should be considered a limitation of the procedure.

#### Web Appendix S5: Subgraphs for visualization

To obtain simpler visualizations of subsets of the mechanisms found in the global TMPDAGs, we construct subgraphs over a subset of variables $S = X_1, \dots, X_s$ in the following way:

1. Draw an empty graph (no edges) including only the nodes in $S$.
2. For each pair of variables $(X_i, X_j) \in S$ with $i \neq j$:
    a. If there exists a directed path $X_i \rightarrow \cdots \rightarrow X_j$ (no undirected edges on the path) that does not include other variables in $S$ (except for $X_i$ and $X_j$), draw a directed edge $X_i \rightarrow X_j$.
    b. If (a) is not fulfilled, but there exists an undirected path or partially directed path (i.e., a path that combines directed edges all pointing the same direction and undirected edges) from $X_i$ to $X_j$ that does not include other variables from $S$ (except for $X_i$ and $X_j$), draw an undirected edge: $X_i - X_j$.

Note that this subgraph construction does not represent confounding and hence it is not a proper causal marginalization of the full TMPDAG, which implies that e.g., d-separation cannot be used on the subgraph. However, it can be used to assert whether there exists a causal path between pairs of variables that is not fully mediated via other considered variables in $S$, and hence also what variables in $S$ one can possibly intervene on to affect other variables in $S$.

*Example:* Consider the TMPDAG in Figure S8 (left) assuming that $(X_1, X_2, X_3, X_4)$ belong to tier 1 and $(X_5, X_6)$ belong to tier 2. We construct a subgraph only including the variables $S = (X_1, X_3, X_4, X_6)$ following the procedure outlined above. Figure S8 (right) shows the resulting graph. We see that the direct edge from $X_4 \rightarrow X_6$ has been preserved (as both were in $S$). The directed path $X_3 \rightarrow X_5 \rightarrow X_6$ has been replaced by an edge $X_3 \rightarrow X_6$ in the subgraph, while the undirected path $X_1 - X_2 - X_4$ has been replaced by an undirected edge $X_1 - X_4$. Finally, the partially directed path $X_1 - X_2 \rightarrow X_5 \rightarrow X_6$ has been replaced by an undirected edge $X_1 - X_6$.

In our displayed subgraphs we displayed robust (≥80 %) paths, but these should be interpreted with some caution. The displayed flow of causal information is relative to the chosen variables of interest. For example, adding a variable could result in one less edge. Imagine we create a subgraph over X and Y, and there is a direct effect which is only 45 % robust, but in every other case there is an indirect effect through Z. We would have a subgraph X → Y but adding Z to the variables of interest would result in an empty subgraph. Thus, subgraphs reflect conditional summaries rather than fixed structural relations.

#### Web Appendix S6: Bootstrap repetitions

For the bootstrap analysis, we sampled with replacement from the full study populations for both "boys" and "girls", creating 100 bootstrap pairs of datasets of the same sizes as the full study populations. For each dataset, we handled missing information as described in Web Appendix S2 (with 100 trees for the imputation model) and S3 (with 1000 trees for the random forest probability model). We then tuned TGES to find a penalty term that resulted in a TMPDAG with 299 to 301 edges.

In a sensitivity analysis, we also estimated TMPDAGs only using complete participants. We compare these results to the main analysis TMPDAGs by computing Structural Hamming Distances (SHDs) separately for boys and girls. Moreover, for each sex, we test whether the TMPDAG from the sensitivity analysis is statistically significantly different from the TMPDAG from the main analysis. We do so by comparing the observed SHD ($\mathrm{SHD}_{\mathrm{complete\ vs.\ analysis}}$) with a distribution of SHDs obtained by comparing each of the bootstrapped TMPDAGs with the main analysis TMPDAG. We report a one-sided *p*-value counting the proportion of bootstraps that result in a larger value of SHD than $\mathrm{SHD}_{\mathrm{complete\ vs.\ analysis}}$.

#### Web Appendix S7: Permutation test

We created 100 copies of the datasets with sex randomly reassigned in each copy, thus breaking any association between sex and all other variables, and consequently also between the causal structure and sex. For each modified dataset, we handled the missing information as described in Web Appendix S2 (with 100 trees for the imputation model) and S3 (with 1000 trees for the random forest probability model) and applied TGES separately for "boys" and "girls" and computed the SHD. Finally, we compared this distribution of SHDs (corresponding to a null hypothesis of no difference in mechanisms for boys and girls) with the observed SHD for the actual comparison of boys and girls and summarized the result as a one-sided *p*-value.

### Exclusion criteria

Exclusion criteria included individuals with congenital anomalies diagnosed before 1 year of age (ICD-10 codes Q00-Q07, Q10-Q18, Q20-Q26, Q300, Q32-Q34, Q35-Q45, Q790, Q792-Q793, Q795, Q60-Q64, Q794, Q65-Q74, Q7402, Q77, Q7800, Q782-Q788, Q750, Q893-Q894, Q80-Q82, Q8726, Q0435, Q411-Q412, Q418, Q710, Q712-Q713, Q720, Q722-Q723, Q730, Q793, Q795, Q7980, Q7982, Q8706, Q206, Q240, Q3381, Q890, Q893, Q86, P350, P351, P371, Q860, Q8680, P350-P351, P371, Q4471, Q6190, Q7484, Q751, Q754, Q7581, Q87, Q936, D821, Q90-Q93, Q96-Q99); individuals with conditions affecting the central nervous system diagnosed before 7 years of age (ICD-10 codes G10-G14, G35-G37, G80-G87, G91), individuals with global developmental delay diagnosis (ICD-10 codes F70, F71-F73, F78, F79); individuals with endocrine and metabolic diseases (ICD-10 codes E00-E07, E20.1, E22.0, E 24, E70, E71, E72, E74, E75, E76, E77, E78, E79, E80, E84, [-E80.4], E85); individuals born before gestational week

28; individuals born with birth weight <1500 g; individuals without a registered father, individual with a dead parent before age 18.

| Variable | Tier | Definition | Classification |
|---|---|---|---|
| **1. WEIGHT** | | | |
| **Weight for gestational age** | At birth | Size for gestational age according to the Intrauterine growth curves by Marsal et al (Acta Pediatr. 1996: 85; 843-8) | Small for gestational age: birth weight < mean birthweight – 2 SD<br>Average for gestational age: mean birth weight – 2SD ≤ birth weight ≤ mean birth weight + 2 SD<br>Large for gestational age: birth weight > mean birthweight + 2 SD |
| **Weight** | Childhood, early adolescence | Age- and sex-specific cutoff points of body mass index (BMI) according to IOTF. | Underweight<br>Normal weight<br>Overweight<br>Obese |
| **Weight** | Late adolescence | Cutoff points of body mass index (BMI) according to WHO | Underweight: BMI below 18.5<br>Normal weight: BMI 18.5-24.9<br>Overweight: BMI 25.0-29.9<br>Obese: BMI above 30.0 |
| **Weight** | Infancy | Weight categories based on BMI z-scores using the WHO Child Growth Standards 2006. | Underweight: z-score below -2<br>Normal weight: z-score between -2 (incl.) and 2 (incl.)<br>Overweight: z-score between 2 (excl.) and 3 (incl.)<br>Obese: z-score above 3 |
| **Maternal weight** | Pregnancy, infancy, childhood, early adolescence | Cutoff points of body mass index (BMI) according to WHO | Underweight: BMI below 18.5<br>Normal weight: BMI 18.5-24.9<br>Overweight: BMI 25.0-29.9<br>Obese: BMI above 30.0 |
| **Paternal weight** | Infancy, childhood, early adolescence | Cutoff points of body mass index (BMI) according to WHO | Underweight: BMI below 18.5<br>Normal weight: BMI 18.5-24.9<br>Overweight: BMI 25.0-29.9<br>Obese: BMI above 30.0 |
| **Gestational weight gain** | At birth | From Harpsøe MC, Basit S, Bager P, et al[4]. | <5kg<br>5-9 kg<br>10-15 kg<br>16-19 kg<br>20-24 kg<br>≥ 25 kg |
| **2. MENTAL HEALTH** | | | |
| **Prosocial problems** | Childhood, early adolescence | Prosocial subscale of parent-reported SDQ.<br>Cutoff is sex specific. | SDQ prosocial score not in bottom 10th percentile<br>SDQ prosocial score in bottom 10th percentile |
| **Prosocial problems** | Late adolescence | Prosocial subscale of child-reported SDQ.<br>Cutoff is sex specific. | SDQ prosocial score not in bottom 10th percentile<br>SDQ prosocial score in bottom 10th percentile |
| **Internalizing problems** | Childhood, early adolescence | Composite scale combining the emotional and peer problems subscales of parent-reported SDQ. Cutoff is sex specific. | SDQ internalizing score not in top 10th percentile<br>SDQ internalizing score in top 10th percentile |
| **Internalizing problems** | Late adolescence | Composite scale combining the emotional and peer problems subscales of child-reported SDQ. Cutoff is sex specific. | SDQ internalizing score not in top 10th percentile<br>SDQ internalizing score in bottom 10th percentile |
| **Externalizing problems** | Childhood, early adolescence | Composite scale combining hyperactivity and behavioral problems subscales of | SDQ externalizing score not in top 10th percentile |

| Variable | Tier | Definition | Classification |
|---|---|---|---|
| | | parent-reported SDQ. Cutoff is sex specific. | SDQ externalizing score in top 10th percentile |
| **Externalizing problems** | Late adolescence | Composite scale combining hyperactivity and behavioral problems subscales of child-reported SDQ. Cutoff is sex specific. | SDQ externalizing score not in top 10th percentile<br>SDQ externalizing score in top 10th percentile |
| **Depression (DAWBA[5])** | Early adolescence | Equals 1 if answered yes to the questions E112, E113_1, E113_2 **. If answered no, or did not answer any of the questions, then 0. | No depression<br>Depression |
| **Stress in children (SiC[6])** | Early adolescence | Global Mean Score (GMS) of the SiC[6] questionnaire<br>GMS less than 2.0: no stress<br>GMS 2.0-2.5 (excl.): moderate stress<br>GMS 2.5 or above: high stress | No stress<br>Moderate stress<br>High stress |
| **Life satisfaction (Cantril[7])** | Early adolescence, late adolescence | Cutoff points of Cantril ladder:<br>0-5: low quality of life<br>6-8: medium quality of life<br>9-10: high quality of life | Low quality of life<br>Medium quality of life<br>High quality of life |
| **Mental well-being (SWEMWBS[8])** | Late adolescence | Cutoff points of SWEMWBS with metric conversion:<br><18.0: probable anxiety or depression<br>18.0-20.0: medium possible anxiety or depression<br>> 20.0: good mental well-being | Probable anxiety or depression<br>Medium possible anxiety or depression<br>Good mental well-being |
| **Body dissatisfaction** | Early adolescence | Defined from DNBC questions E125_1, E125_2, E126_1 and E126_2[9] and based on The Figure Rating Scale by Collins[10]. | None: No body dissatisfaction (score 0)<br>Low: Low level of body dissatisfaction (score -1 or 1)<br>High: High level of body dissatisfaction (score from -2 to -6 or from 2 to 6). |
| **Major Depression Inventory score** | Late adolescence | Cutoff points of MDI score:<br>< 20: no depressive symptoms<br>21-25: mild depression<br>26-30: moderate depression<br>>30: severe depression | None: No depressive symptoms<br>Mild: Mild depression<br>Moderate: Moderate depression<br>Severe: Severe depression |
| **Neurodevelopmental diagnosis*** ** | Childhood, early adolescence, late adolescence | ICD-10 codes F80–F84, F88–F90, F95, F98.8 | No diagnosis<br>One or more diagnoses |
| **Affective disorders** *** | Childhood, early adolescence, late adolescence | ICD-10 codes F30–F39, F40–F48, F93 | No diagnosis<br>One or more diagnoses |
| **Psychotic disorders** *** | Late adolescence | ICD-10 codes F20-F29 | No diagnosis<br>One or more diagnoses |
| **Other psychiatric diagnoses*** ** | Childhood, early adolescence, late adolescence | ICD-10 codes F50, F51, F91, F94, F98.0–98.7, F98.9, F99, F60–F69. | No diagnosis<br>One or more diagnoses |

| | | | |
|---|---|---|---|
| **3. LIFESTYLE RISK FACTORS** | | | |
| **Physical activity** | Childhood | Parents answer to “How many hours is your daughter physically active in kindergarten, school or at the leisure centre/school leisure centre?”: | Never or very rarely<br>Less than 1 hour a week<br>1-2 hours a week<br>3-4 hours a week<br>5-6 hours a week<br>6 hours or more |
| **Physical activity** | Early adolescence | Indicators for daily activity and leisure time activity based on WHO’s definition of physical activity among children and young people[11]. | Inactive<br>Lightly active<br>Moderately active<br>Vigorously active |
| **Screen time** | Early adolescence | Total (minimum) number of screen time hours per day (irrespective of type). We assign weights to the estimates of weekdays (5/7) and weekend days (2/7) | 2 or less hours<br>2 to 4 hours<br>4 to 6 hours<br>6 or more hours |
| **Sleep time** | Early adolescence | Time participants go to sleep in the weekdays | 19:00 or earlier<br>19:30 to 20:00<br>20:30 to 21:00<br>21:30 to 22:00<br>22:30 to 23:00<br>23:30 or later |
| **Sleep time** | Late adolescence | Time the participants go to sleep in the weekdays | 20:00 or earlier<br>21:00 to 22:00<br>23:00 to 00:00<br>01:00 to 02:00<br>03:00 or later |
| **4. OTHER** | | | |
| **Gestational age** | At birth | Units are completed weeks | 28-31weeks<br>32-36 weeks<br>37-39 weeks<br>40-42 weeks<br>Over 42 weeks |
| **Full-time breastfeeding** | Infancy | | Never full-time breastfeeding<br>Full-time breastfeeding up to 4 months<br>Full-time breastfeeding 4-6 months<br>Full-time breastfeeding more than 6 months |
| **Adversity: material** | Infancy, childhood, early adolescence, late adolescence | One count per year of life for each parent being unemployed for at least 12 months. | No occurrence<br>Minimum one occurrence |
| **Adversity: loss** | Infancy, childhood, early adolescence, late adolescence | If death: One count in the year a liveborn sibling or half sibling dies.<br><br>If parental illness: One count per year of life a parent gets diagnosed with one of the illnesses related to the mortality included in the Charlson comorbidity index. All diagnosis codes listed in Thygesen SK, Christiansen CF, Christensen S, et al. 2011[12]. Primary and | No occurrence<br>Minimum one occurrence |

| | | | |
|---|---|---|---|
| | | secondary diagnosis are included, and all hospital contacts are counted as an event. In DANLIFE, all hospital contacts are also included from 1995 and onwards (ICD-10).<br><br>If sibling or half sibling illness: One count per year of life for each sibling diagnosed with one of the seven most common somatic illnesses related to mortality in children aged 0–18 years in Denmark (ICD-10: C00-C96, Q20-Q28, Q00-Q07, G80-G83, G40-G41, I42-I43, E75). Primary and secondary diagnosis are included, and all hospital contacts are counted as an event. | |
| **Adversity: family** | Infancy, childhood, early adolescence, late adolescence | If foster: One count per year of life overlapping with a calendar year in which the child was registered as placed in out-of-home care.<br><br>If parental psychiatric illness: hospital contacts (incl. admission and outpatient) with a diagnosis related to psychiatric illness (excluding main diagnoses related to alcohol and drug abuse). All F-diagnosis are included except diagnosis related to alcohol or drug abuse (same diagnosis codes used to define the variables parental alcohol abuse and parental drug abuse). Diagnosis code for tobacco abuse is also excluded. Primary and secondary diagnosis are included, and all hospital contacts are counted as an event.<br><br>If sibling psychiatric illness: One count per year of life for each sibling with a hospital admission with a diagnosis related to psychiatric illness. All F-diagnosis are included. No diagnoses are excluded. Primary and secondary diagnosis are included, and all hospital contacts are counted as an event. Half siblings are also included.<br><br>If parental alcohol abuse: One count per year of life for each parent diagnosed with an illness related to alcohol abuse or receiving a prescription of a drug used in treatment of alcohol addiction. ICD-10: F10, F70, G621, I1426, K852, K860, K292, N07BB (N07BB01 N07BB2 N07BB3 N07BB4 N07BB5).<br><br>If parental drug abuse: One count per year of life for each parent diagnosed with an illness related to drug abuse or receiving a prescription of drugs used in | No occurrence<br>Minimum one occurrence |

| | | | |
|---|---|---|---|
| | | treatment of drug addiction. ICD-10: F11-F16, F18, F19N07BC (N07BC01, N07BC02, N07BC03, N07BC04, N07BC05, N07BC06, N07BC51).<br><br>If maternal separation: One count per year of life overlapping with a calendar year in which the mother was separated from a partner. | |
| **Household education** | Pregnancy | Highest level of completed or ongoing education in the household at the time of the child's birth, from Danish education registries (DISCED-15*). | Low: DISCED 10, 20*<br>Medium: DISCED 30, 40*<br>High: DISCED 50, 60, 70, 80* |
| **Household income** | At birth, infancy, childhood, early adolescence, late adolescence | Equalized household income accounting for size and composition, from the Income Statistics Register. Categorized into quartiles. | Q1<br>Q2<br>Q3<br>Q4 |
| **Sex** | At birth | Biological sex of child from DNBC, validated via the Civil Registration System (CPR). | Male<br>Female |
| **Parity** | At birth | Number of previous pregnancies, from the Danish Medical Birth Register. | No previous pregnancies<br>One<br>Two<br>Three or more |
| **Maternal age** | Pregnancy | Age of the mother a year before the time of the child's birth. The intervals are made such that they correspond to what is traditionally done in the literature at age at time of birth, i.e. <25, 25-29. | <24 years<br>24–28 years<br>29–33 years<br>≥34 years |
| **Paternal age** | Pregnancy | Age of the father a year before the time of the child's birth. The intervals are made such that they correspond to what is traditionally done in the literature at age at time of birth, i.e. <25, 25-29. | <24 years<br>24–28 years<br>29–33 years<br>≥34 years |
| **Maternal smoking** | Pregnancy | Smoking status during pregnancy from the Danish Medical Birth Register. | Did not smoke<br>Quit during pregnancy<br>Smoked during pregnancy |

*Table S1: Categorization and description of measurements included in the data analysis. Tiers correspond to the age of the child as follows: Pregnancy (16 weeks of gestation -interview 1-; 30 weeks of gestation -interview 2-), At birth, Infancy (6 months postpartum -interview 3-); 18 months postpartum -interview 4-), Childhood (7 years), Early adolescence (11 years), Mid Adolescence (14 years), Late Adolescence (18 years)*

**DISCED refers to the Danish International Standard Classification of Education (Classification on Education (DISCED-15), which is Denmark's national adaptation of UNESCO's International Standard Classification of Education (ISCED). This variable is categorized as Low Education (DISCED 10, 20); Medium Education (DISCED 30, 40) High Education (DISCED 50, 60, 70, 80).*

***[9] E112 During the last 4 weeks, have there been times when you have been very sad,* miserable, unhappy or tearful? 1. Yes; 2. No; 99. not answered

E113_1 During the last 4 weeks, has there been a period when you have been really miserable nearly every day? 1. Yes; 2. No; 99. not answered; 100. not applicable

E113_2 During the time when you have been miserable, have you been really miserable for most of the day? (dvs. most hours, where you felt miserable) 1. Yes; 2. No; 99. not answered; 100. not applicable

******* *These variables capture the presence of psychiatric diagnoses based on diagnostic records from the Danish National Patient Register (Landspatientregisteret LPR). Diagnoses were extracted from all inpatient admissions and outpatient contacts.*

Abbreviations: BMI: Body Mass Index, ICD-10: 10th revision of the International Classification of Diseases; SDQ: strengths and difficulties questionnaire; DAWBA: Development and Well-Being Assessment; SiC: Stress in Children Questionnaire; SWEMWBS: Short Warwick-Edinburgh Well-Being Score

| Tier | Variable | Level | Female study population | Male study population | Female complete participants | Male complete participants | Female analysis sample | Male analysis sample |
|---|---|---|---|---|---|---|---|---|
| **Pregnancy** | Maternal Weight | BMI below 18.5 | 1426 (0.04) | 1541 (0.04) | 279 (0.04) | 209 (0.04) | 1482 (0.05) | 1467 (0.04) |
| | | BMI 18.5 - 24.9 | 21842 (0.67) | 23118 (0.66) | 4877 (0.70) | 3666 (0.71) | 22377 (0.68) | 24164 (0.69) |
| | | BMI 25.0 - 29.9 | 6300 (0.19) | 6740 (0.19) | 1255 (0.18) | 925 (0.18) | 6364 (0.19) | 6797 (0.20) |
| | | BMI above 30.0 | 2709 (0.08) | 2817 (0.08) | 470 (0.07) | 310 (0.06) | 2589 (0.08) | 2353 (0.07) |
| | | Missing | 535 (0.02) | 565 (0.02) | 108 (0.02) | 71 (0.01) | | |
| | Paternal age | <24 years | 1102 (0.03) | 1207 (0.03) | 140 (0.02) | 98 (0.02) | 958 (0.03) | 1029 (0.03) |
| | | 24-28 years | 8060 (0.25) | 8473 (0.24) | 1607 (0.23) | 1206 (0.23) | 7883 (0.24) | 8411 (0.24) |
| | | 29-33 years | 13033 (0.40) | 13954 (0.40) | 2825 (0.40) | 2053 (0.40) | 12849 (0.39) | 13635 (0.39) |
| | | Above 34 years | 10609 (0.32) | 11138 (0.32) | 2417 (0.35) | 1824 (0.35) | 11122 (0.34) | 11706 (0.34) |
| | | Missing | 8 (0.00) | 9 (0.00) | | | | |
| | Maternal age | <24 years | 2504 (0.08) | 2728 (0.08) | 338 (0.05) | 236 (0.05) | 2154 (0.07) | 2239 (0.06) |
| | | 24-28 years | 11815 (0.36) | 12517 (0.36) | 2437 (0.35) | 1779 (0.34) | 11655 (0.36) | 12146 (0.35) |
| | | 29-33 years | 12866 (0.39) | 13562 (0.39) | 2855 (0.41) | 2153 (0.42) | 12939 (0.39) | 13930 (0.40) |
| | | Above 34 years | 5627 (0.17) | 5974 (0.17) | 1359 (0.19) | 1013 (0.20) | 6064 (0.18) | 6466 (0.19) |
| | | Missing | | | | | | |
| | Household education | Low: DISCED 10, 20 | 1306 (0.04) | 1521 (0.04) | 132 (0.02) | 80 (0.02) | 962 (0.03) | 857 (0.02) |
| | | Medium: DISCED 30, 40 | 13270 (0.40) | 13932 (0.40) | 2312 (0.33) | 1619 (0.31) | 12601 (0.38) | 12775 (0.37) |
| | | High: DISCED 50, 60, 70, 80 | 18236 (0.56) | 19328 (0.56) | 4545 (0.65) | 3482 (0.67) | 19249 (0.59) | 21149 (0.61) |
| | | Missing | | | | | | |
| | Maternal smoking | No smoking | 24236 (0.74) | 25873 (0.74) | 5525 (0.79) | 4241 (0.82) | 24862 (0.76) | 27135 (0.78) |
| | | Quit smoking | 3130 (0.10) | 3257 (0.09) | 630 (0.09) | 402 (0.08) | 3161 (0.10) | 2968 (0.09) |

| Tier | Variable | Level | Female study population | Male study population | Female complete participants | Male complete participants | Female analysis sample | Male analysis sample |
|---|---|---|---|---|---|---|---|---|
| | | Smoked < 10 cigarettes/day | 2958 (0.09) | 3068 (0.09) | 501 (0.07) | 318 (0.06) | 2683 (0.08) | 2653 (0.08) |
| | | Smoked > 9 cigarettes/day | 2457 (0.07) | 2540 (0.07) | 327 (0.05) | 212 (0.04) | 2106 (0.06) | 2025 (0.06) |
| | | Missing | 31 (0.00) | 43 (0.00) | 6 (0.00) | 8 (0.00) | | |
| **At birth** | Gestational age | 28-31 weeks | 41 (0.00) | 68 (0.00) | 8 (0.00) | 6 (0.00) | 67 (0.00) | 42 (0.00) |
| | | 32-36 weeks | 1135 (0.03) | 1334 (0.04) | 206 (0.03) | 190 (0.04) | 1101 (0.03) | 1369 (0.04) |
| | | 37-39 weeks | 12081 (0.37) | 12994 (0.37) | 2557 (0.37) | 1913 (0.37) | 12065 (0.37) | 12758 (0.37) |
| | | 40-42 weeks | 19424 (0.59) | 20252 (0.58) | 4188 (0.60) | 3058 (0.59) | 19429 (0.59) | 20500 (0.59) |
| | | Over 42 weeks | 131 (0.00) | 133 (0.00) | 30 (0.00) | 14 (0.00) | 150 (0.00) | 112 (0.00) |
| | | Missing | | | | | | |
| | Gestational weight gain | <5 kg | 590 (0.02) | 535 (0.02) | 115 (0.02) | 63 (0.01) | 721 (0.02) | 514 (0.01) |
| | | 5-9 kg | 2864 (0.09) | 2741 (0.08) | 724 (0.10) | 498 (0.10) | 3575 (0.11) | 3476 (0.10) |
| | | 10-15 kg | 11539 (0.35) | 11956 (0.34) | 3361 (0.48) | 2449 (0.47) | 15554 (0.47) | 16106 (0.46) |
| | | 16-19 kg | 5132 (0.16) | 5651 (0.16) | 1430 (0.20) | 1085 (0.21) | 6519 (0.20) | 7299 (0.21) |
| | | 20-24 kg | 3558 (0.11) | 4002 (0.12) | 876 (0.13) | 736 (0.14) | 4174 (0.13) | 5050 (0.15) |
| | | >=25 kg | 1780 (0.05) | 2071 (0.06) | 389 (0.06) | 277 (0.05) | 2269 (0.07) | 2336 (0.07) |
| | | Missing | 7349 (0.22) | 7825 (0.22) | 94 (0.01) | 73 (0.01) | | |
| | Household income | Q1 | 4576 (0.14) | 4866 (0.14) | 743 (0.11) | 537 (0.10) | 4182 (0.13) | 4201 (0.12) |
| | | Q2 | 8155 (0.25) | 8690 (0.25) | 1613 (0.23) | 1170 (0.23) | 7922 (0.24) | 8398 (0.24) |
| | | Q3 | 9772 (0.30) | 10257 (0.29) | 2173 (0.31) | 1584 (0.31) | 9874 (0.30) | 10414 (0.30) |
| | | Q4 | 10289 (0.31) | 10958 (0.32) | 2453 (0.35) | 1890 (0.36) | 10834 (0.33) | 11768 (0.34) |
| | | Missing | 20 (0.00) | 10 (0.00) | 7 (0.00) | | | |
| | Parity | No previous pregnancies | 15312 (0.47) | 16262 (0.47) | 3298 (0.47) | 2476 (0.48) | 15538 (0.47) | 16623 (0.48) |

| Tier | Variable | Level | Female study population | Male study population | Female complete participants | Male complete participants | Female analysis sample | Male analysis sample |
|---|---|---|---|---|---|---|---|---|
| | | One | 12224 (0.37) | 12970 (0.37) | 2581 (0.37) | 1862 (0.36) | 11958 (0.36) | 12651 (0.36) |
| | | Two | 4306 (0.13) | 4493 (0.13) | 942 (0.13) | 720 (0.14) | 4435 (0.14) | 4638 (0.13) |
| | | Three or more | 951 (0.03) | 1034 (0.03) | 168 (0.02) | 123 (0.02) | 881 (0.03) | 869 (0.02) |
| | | Missing | 19 (0.00) | 22 (0.00) | | | | |
| | Weight for gestational age | Small for gestational age | 717 (0.02) | 745 (0.02) | 121 (0.02) | 96 (0.02) | 633 (0.02) | 656 (0.02) |
| | | Average for gestational age | 30813 (0.94) | 32528 (0.94) | 6585 (0.94) | 4865 (0.94) | 30825 (0.94) | 32700 (0.94) |
| | | Large for gestational age | 1274 (0.04) | 1497 (0.04) | 283 (0.04) | 220 (0.04) | 1354 (0.04) | 1425 (0.04) |
| | | Missing | 8 (0.00) | 11 (0.00) | | | | |
| **Infancy** | Paternal weight | BMI below 18.5 | 100 (0.00) | 104 (0.00) | 28 (0.00) | 26 (0.01) | 128 (0.00) | 171 (0.00) |
| | | BMI 18.5 - 24.9 | 12547 (0.38) | 13099 (0.38) | 3791 (0.54) | 2813 (0.54) | 18029 (0.55) | 19035 (0.55) |
| | | BMI 25.0 - 29.9 | 9025 (0.28) | 9694 (0.28) | 2523 (0.36) | 1891 (0.36) | 12402 (0.38) | 13414 (0.39) |
| | | BMI above 30.0 | 1675 (0.05) | 1629 (0.05) | 430 (0.06) | 282 (0.05) | 2253 (0.07) | 2161 (0.06) |
| | | Missing | 9465 (0.29) | 10255 (0.29) | 217 (0.03) | 169 (0.03) | | |
| | Maternal weight | BMI below 18.5 | 929 (0.03) | 969 (0.03) | 248 (0.04) | 186 (0.04) | 1353 (0.04) | 1450 (0.04) |
| | | BMI 18.5 - 24.9 | 14324 (0.44) | 15000 (0.43) | 4256 (0.61) | 3159 (0.61) | 22035 (0.67) | 23621 (0.68) |
| | | BMI 25.0 - 29.9 | 4477 (0.14) | 4757 (0.14) | 1221 (0.17) | 898 (0.17) | 6997 (0.21) | 7304 (0.21) |
| | | BMI above 30.0 | 1707 (0.05) | 1796 (0.05) | 413 (0.06) | 268 (0.05) | 2427 (0.07) | 2406 (0.07) |
| | | Missing | 11375 (0.35) | 12259 (0.35) | 851 (0.12) | 670 (0.13) | | |
| | Weight | Z-score below -2 | 319 (0.01) | 544 (0.02) | 103 (0.01) | 118 (0.02) | 470 (0.01) | 793 (0.02) |
| | | Z-score between -2 (incl.) and 2 (incl.) | 17990 (0.55) | 18158 (0.52) | 5449 (0.78) | 3933 (0.76) | 31105 (0.95) | 32231 (0.93) |
| | | Z-score between 2 | 812 (0.02) | 1101 (0.03) | 232 (0.03) | 227 (0.04) | 1098 (0.03) | 1530 (0.04) |

| Tier | Variable | Level | Female study population | Male study population | Female complete participants | Male complete participants | Female analysis sample | Male analysis sample |
|---|---|---|---|---|---|---|---|---|
| | | (excl.) and 3 (incl.) | | | | | | |
| | | Z-score above 3 | 91 (0.00) | 198 (0.01) | 28 (0.00) | 34 (0.01) | 139 (0.00) | 227 (0.01) |
| | | Missing | 13600 (0.41) | 14780 (0.42) | 1177 (0.17) | 869 (0.17) | | |
| | Adversity: family | No occurrence | 30581 (0.93) | 32368 (0.93) | 6725 (0.96) | 4997 (0.96) | 31026 (0.95) | 33076 (0.95) |
| | | Minimum one occurrence | 2231 (0.07) | 2413 (0.07) | 264 (0.04) | 184 (0.04) | 1786 (0.05) | 1705 (0.05) |
| | Adversity: loss | No occurrence | 31975 (0.97) | 33871 (0.97) | 6833 (0.98) | 5042 (0.97) | 31997 (0.98) | 33773 (0.97) |
| | | Minimum one occurrence | 837 (0.03) | 910 (0.03) | 156 (0.02) | 139 (0.03) | 815 (0.02) | 1008 (0.03) |
| | Adversity: material | No occurrence | 31395 (0.96) | 33336 (0.96) | 6746 (0.97) | 5015 (0.97) | 31481 (0.96) | 33455 (0.96) |
| | | Minimum one occurrence | 1417 (0.04) | 1445 (0.04) | 243 (0.03) | 166 (0.03) | 1331 (0.04) | 1326 (0.04) |
| | Full-time breastfeeding | Never full-time breastfeeding | 959 (0.03) | 1133 (0.03) | 236 (0.03) | 195 (0.04) | 1207 (0.04) | 1381 (0.04) |
| | | Full-time breastfeeding up to 4 months | 11854 (0.36) | 13269 (0.38) | 2954 (0.42) | 2250 (0.43) | 14384 (0.44) | 15842 (0.46) |
| | | Full-time breastfeeding 4-6 months | 9722 (0.30) | 9564 (0.27) | 2859 (0.41) | 1970 (0.38) | 12746 (0.39) | 12544 (0.36) |
| | | Full-time breastfeeding more than 6 months | 3331 (0.10) | 3418 (0.10) | 940 (0.13) | 766 (0.15) | 4475 (0.14) | 5014 (0.14) |
| | | Missing | 6946 (0.21) | 7397 (0.21) | | | | |
| | Household income | Q1 | 4530 (0.14) | 4816 (0.14) | 704 (0.10) | 503 (0.10) | 4151 (0.13) | 4267 (0.12) |
| | | Q2 | 8002 (0.24) | 8430 (0.24) | 1558 (0.22) | 1136 (0.22) | 7709 (0.23) | 8087 (0.23) |
| | | Q3 | 9712 (0.30) | 10371 (0.30) | 2221 (0.32) | 1609 (0.31) | 10018 (0.31) | 10519 (0.30) |
| | | Q4 | 10563 (0.32) | 11151 (0.32) | 2506 (0.36) | 1933 (0.37) | 10934 (0.33) | 11908 (0.34) |
| | | Missing | 5 (0.00) | 13 (0.00) | | | | |

| Tier | Variable | Level | Female study population | Male study population | Female complete participants | Male complete participants | Female analysis sample | Male analysis sample |
|---|---|---|---|---|---|---|---|---|
| **Childhood** | Weight Childhood | Underweight | 2254 (0.07) | 2212 (0.06) | 820 (0.12) | 562 (0.11) | 3849 (0.12) | 3668 (0.11) |
| | | Normal weight | 14689 (0.45) | 16346 (0.47) | 5239 (0.75) | 4052 (0.78) | 25793 (0.79) | 28541 (0.82) |
| | | Overweight | 1648 (0.05) | 1440 (0.04) | 513 (0.07) | 310 (0.06) | 2715 (0.08) | 2248 (0.06) |
| | | Obese | 272 (0.01) | 205 (0.01) | 70 (0.01) | 41 (0.01) | 455 (0.01) | 324 (0.01) |
| | | Missing | 13949 (0.43) | 14578 (0.42) | 347 (0.05) | 216 (0.04) | | |
| | Paternal weight | BMI below 18.5 | 41 (0.00) | 56 (0.00) | 12 (0.00) | 18 (0.00) | 61 (0.00) | 124 (0.00) |
| | | BMI 18.5 - 24.9 | 8295 (0.25) | 8885 (0.26) | 2994 (0.43) | 2279 (0.44) | 14751 (0.45) | 15787 (0.45) |
| | | BMI 25.0 - 29.9 | 8153 (0.25) | 8600 (0.25) | 2891 (0.41) | 2132 (0.41) | 14752 (0.45) | 15501 (0.45) |
| | | BMI above 30.0 | 1837 (0.06) | 1987 (0.06) | 622 (0.09) | 444 (0.09) | 3248 (0.10) | 3369 (0.10) |
| | | Missing | 14486 (0.44) | 15253 (0.44) | 470 (0.07) | 308 (0.06) | | |
| | Maternal weight | BMI below 18.5 | 423 (0.01) | 478 (0.01) | 143 (0.02) | 108 (0.02) | 774 (0.02) | 745 (0.02) |
| | | BMI 18.5 - 24.9 | 12761 (0.39) | 13593 (0.39) | 4502 (0.64) | 3362 (0.65) | 20699 (0.63) | 22248 (0.64) |
| | | BMI 25.0 - 29.9 | 4623 (0.14) | 4798 (0.14) | 1586 (0.23) | 1152 (0.22) | 7930 (0.24) | 8178 (0.24) |
| | | BMI above 30.0 | 1987 (0.06) | 2113 (0.06) | 635 (0.09) | 462 (0.09) | 3409 (0.10) | 3610 (0.10) |
| | | Missing | 13018 (0.40) | 13799 (0.40) | 123 (0.02) | 97 (0.02) | | |
| | Adversity: family | No occurrence | 24912 (0.76) | 26530 (0.76) | 5742 (0.82) | 4380 (0.85) | 25716 (0.78) | 27989 (0.80) |
| | | Minimum one occurrence | 7900 (0.24) | 8251 (0.24) | 1247 (0.18) | 801 (0.15) | 7096 (0.22) | 6792 (0.20) |
| | Adversity: loss | No occurrence | 30060 (0.92) | 31873 (0.92) | 6436 (0.92) | 4804 (0.93) | 29934 (0.91) | 32114 (0.92) |
| | | Minimum one occurrence | 2752 (0.08) | 2908 (0.08) | 553 (0.08) | 377 (0.07) | 2878 (0.09) | 2667 (0.08) |
| | Adversity: material | No occurrence | 29385 (0.90) | 31157 (0.90) | 6410 (0.92) | 4777 (0.92) | 29665 (0.90) | 31522 (0.91) |
| | | Minimum one occurrence | 3427 (0.10) | 3624 (0.10) | 579 (0.08) | 404 (0.08) | 3147 (0.10) | 3259 (0.09) |

| Tier | Variable | Level | Female study population | Male study population | Female complete participants | Male complete participants | Female analysis sample | Male analysis sample |
|---|---|---|---|---|---|---|---|---|
| | Household income | Q1 | 4838 (0.15) | 5186 (0.15) | 683 (0.10) | 491 (0.09) | 4228 (0.13) | 4315 (0.12) |
| | | Q2 | 8032 (0.24) | 8623 (0.25) | 1550 (0.22) | 1113 (0.21) | 7917 (0.24) | 8280 (0.24) |
| | | Q3 | 9648 (0.29) | 10270 (0.30) | 2269 (0.32) | 1687 (0.33) | 9881 (0.30) | 10730 (0.31) |
| | | Q4 | 10288 (0.31) | 10697 (0.31) | 2487 (0.36) | 1890 (0.36) | 10786 (0.33) | 11456 (0.33) |
| | | Missing | 6 (0.00) | 5 (0.00) | | | | |
| | Externalizing problems | SDQ externalizing score not in top 10th percentile | 18129 (0.55) | 18857 (0.54) | 6346 (0.91) | 4687 (0.90) | 29385 (0.90) | 31028 (0.89) |
| | | SDQ externalizing score in top 10th percentile | 2158 (0.07) | 2687 (0.08) | 607 (0.09) | 475 (0.09) | 3427 (0.10) | 3753 (0.11) |
| | | Missing | 12525 (0.38) | 13237 (0.38) | 36 (0.01) | 19 (0.00) | | |
| | Internalizing problems | SDQ internalizing score not in top 10th percentile | 17287 (0.53) | 19189 (0.55) | 6027 (0.86) | 4667 (0.90) | 27844 (0.85) | 31017 (0.89) |
| | | SDQ internalizing score in top 10th percentile | 3058 (0.09) | 2409 (0.07) | 949 (0.14) | 506 (0.10) | 4968 (0.15) | 3764 (0.11) |
| | | Missing | 12467 (0.38) | 13183 (0.38) | 13 (0.00) | 8 (0.00) | | |
| | Prosocial problems | SDQ prosocial score not in bottom 10th percentile | 16661 (0.51) | 17904 (0.51) | 5756 (0.82) | 4326 (0.83) | 26877 (0.82) | 29088 (0.84) |
| | | SDQ prosocial score in bottom 10th percentile | 3690 (0.11) | 3703 (0.11) | 1222 (0.17) | 848 (0.16) | 5935 (0.18) | 5693 (0.16) |
| | | Missing | 12461 (0.38) | 13174 (0.38) | 11 (0.00) | 7 (0.00) | | |
| | Physical activity | Never or very rarely | 33 (0.00) | 6 (0.00) | 13 (0.00) | | 52 (0.00) | 28 (0.00) |
| | | Less than 1 hour a week | | 15 (0.00) | | 123 (0.02) | | |
| | | 1-2 hours a week | 580 (0.02) | 471 (0.01) | 199 (0.03) | | 929 (0.03) | 781 (0.02) |
| | | 3-4 hours a week | 3594 (0.11) | 2644 (0.08) | 1255 (0.18) | 684 (0.13) | 5856 (0.18) | 4445 (0.13) |

| Tier | Variable | Level | Female study population | Male study population | Female complete participants | Male complete participants | Female analysis sample | Male analysis sample |
|---|---|---|---|---|---|---|---|---|
| | | 5-6 hours a week | 5833 (0.18) | 5316 (0.15) | 2044 (0.29) | 1281 (0.25) | 9412 (0.29) | 8542 (0.25) |
| | | 6 hours or more | 10256 (0.31) | 13078 (0.38) | 3458 (0.49) | 3072 (0.59) | 16563 (0.50) | 20985 (0.60) |
| | | Missing | 12516 (0.38) | 13251 (0.38) | 20 (0.00) | 21 (0.00) | | |
| | Neurodevelopmental | No diagnosis | 32562 (0.99) | 33827 (0.97) | 6953 (0.99) | 5086 (0.98) | 32583 (0.99) | 34024 (0.98) |
| | | One or more diagnoses | 250 (0.01) | 954 (0.03) | 36 (0.01) | 95 (0.02) | 229 (0.01) | 757 (0.02) |
| | Other psychiatric diagnoses | No diagnosis | 32660 (1.00) | 34420 (0.99) | 6960 (1.00) | 5148 (0.99) | 32647 (0.99) | 34519 (0.99) |
| | | One or more diagnoses | 152 (0.00) | 361 (0.01) | 29 (0.00) | 33 (0.01) | 165 (0.01) | 262 (0.01) |
| **Early adolescence** | Weight | Underweight | 3017 (0.09) | 2111 (0.06) | 1258 (0.18) | 653 (0.13) | 5760 (0.18) | 4322 (0.12) |
| | | Normal weight | 11660 (0.36) | 12338 (0.35) | 4818 (0.69) | 3829 (0.74) | 24325 (0.74) | 27208 (0.78) |
| | | Overweight | 1186 (0.04) | 1277 (0.04) | 450 (0.06) | 367 (0.07) | 2524 (0.08) | 2933 (0.08) |
| | | Obese | 105 (0.00) | 128 (0.00) | 31 (0.00) | 36 (0.01) | 203 (0.01) | 318 (0.01) |
| | | Missing | 16844 (0.51) | 18927 (0.54) | 432 (0.06) | 296 (0.06) | | |
| | Paternal weight | BMI below 18.5 | 28 (0.00) | 36 (0.00) | 12 (0.00) | 13 (0.00) | 53 (0.00) | 81 (0.00) |
| | | BMI 18.5 - 24.9 | 6820 (0.21) | 6742 (0.19) | 2848 (0.41) | 2149 (0.41) | 14351 (0.44) | 15327 (0.44) |
| | | BMI 25.0 - 29.9 | 6465 (0.20) | 6652 (0.19) | 2631 (0.38) | 1997 (0.39) | 14825 (0.45) | 15565 (0.45) |
| | | BMI above 30.0 | 1611 (0.05) | 1536 (0.04) | 624 (0.09) | 472 (0.09) | 3583 (0.11) | 3808 (0.11) |
| | | Missing | 17888 (0.55) | 19815 (0.57) | 874 (0.13) | 550 (0.11) | | |
| | Maternal weight | BMI below 18.5 | 275 (0.01) | 299 (0.01) | 109 (0.02) | 90 (0.02) | 568 (0.02) | 621 (0.02) |
| | | BMI 18.5 - 24.9 | 10383 (0.32) | 10399 (0.30) | 4235 (0.61) | 3191 (0.62) | 19650 (0.60) | 21384 (0.61) |

| Tier | Variable | Level | Female study population | Male study population | Female complete participants | Male complete participants | Female analysis sample | Male analysis sample |
|---|---|---|---|---|---|---|---|---|
| | | BMI 25.0 - 29.9 | 4042 (0.12) | 4047 (0.12) | 1643 (0.24) | 1221 (0.24) | 8366 (0.25) | 8616 (0.25) |
| | | BMI above 30.0 | 1816 (0.06) | 1639 (0.05) | 744 (0.11) | 493 (0.10) | 4228 (0.13) | 4160 (0.12) |
| | | Missing | 16296 (0.50) | 18397 (0.53) | 258 (0.04) | 186 (0.04) | | |
| | Depression (DAWBA) | No depression | 17349 (0.53) | 16215 (0.47) | 6646 (0.95) | 4926 (0.95) | 31909 (0.97) | 34447 (0.99) |
| | | Depression | 548 (0.02) | 190 (0.01) | 163 (0.02) | 46 (0.01) | 903 (0.03) | 334 (0.01) |
| | | Missing | 14915 (0.45) | 18376 (0.53) | 180 (0.03) | 209 (0.04) | | |
| | Stress in children (SiC) | No stress | 12662 (0.39) | 11581 (0.33) | 4874 (0.70) | 3558 (0.69) | 22406 (0.68) | 23711 (0.68) |
| | | Moderate stress | 5066 (0.15) | 5062 (0.15) | 1778 (0.25) | 1388 (0.27) | 8836 (0.27) | 9962 (0.29) |
| | | High stress | 780 (0.02) | 543 (0.02) | 263 (0.04) | 143 (0.03) | 1570 (0.05) | 1108 (0.03) |
| | | Missing | 14304 (0.44) | 17595 (0.51) | 74 (0.01) | 92 (0.02) | | |
| | Adversity: family | No occurrence | 26407 (0.80) | 28083 (0.81) | 6012 (0.86) | 4513 (0.87) | 27005 (0.82) | 29055 (0.84) |
| | | Minimum one occurrence | 6405 (0.20) | 6698 (0.19) | 977 (0.14) | 668 (0.13) | 5807 (0.18) | 5726 (0.16) |
| | Adversity: loss | No occurrence | 30342 (0.92) | 32209 (0.93) | 6516 (0.93) | 4815 (0.93) | 30368 (0.93) | 32117 (0.92) |
| | | Minimum one occurrence | 2470 (0.08) | 2572 (0.07) | 473 (0.07) | 366 (0.07) | 2444 (0.07) | 2664 (0.08) |
| | Adversity: material | No occurrence | 31545 (0.96) | 33468 (0.96) | 6815 (0.98) | 5063 (0.98) | 31726 (0.97) | 33684 (0.97) |
| | | Minimum one occurrence | 1267 (0.04) | 1313 (0.04) | 174 (0.02) | 118 (0.02) | 1086 (0.03) | 1097 (0.03) |
| | Body dissatisfaction | High | 854 (0.03) | 843 (0.02) | 281 (0.04) | 232 (0.04) | 1572 (0.05) | 1778 (0.05) |
| | | Low | 5458 (0.17) | 4218 (0.12) | 2004 (0.29) | 1277 (0.25) | 9743 (0.30) | 8961 (0.26) |
| | | None | 11637 (0.35) | 11382 (0.33) | 4550 (0.65) | 3475 (0.67) | 21497 (0.66) | 24042 (0.69) |
| | | Missing | 14863 (0.45) | 18338 (0.53) | 154 (0.02) | 197 (0.04) | | |
| | Life satisfaction (Cantril) | Low quality of life | 1224 (0.04) | 829 (0.02) | 424 (0.06) | 243 (0.05) | 2323 (0.07) | 1790 (0.05) |

| Tier | Variable | Level | Female study population | Male study population | Female complete participants | Male complete participants | Female analysis sample | Male analysis sample |
|---|---|---|---|---|---|---|---|---|
| | | Medium quality of life | 8020 (0.24) | 7368 (0.21) | 2973 (0.43) | 2172 (0.42) | 14101 (0.43) | 14770 (0.42) |
| | | High quality of life | 9300 (0.28) | 9047 (0.26) | 3526 (0.50) | 2680 (0.52) | 16388 (0.50) | 18221 (0.52) |
| | | Missing | 14268 (0.43) | 17537 (0.50) | 66 (0.01) | 86 (0.02) | | |
| | Household income | Q1 | 4899 (0.15) | 5071 (0.15) | 651 (0.09) | 453 (0.09) | 4230 (0.13) | 4081 (0.12) |
| | | Q2 | 8014 (0.24) | 8614 (0.25) | 1555 (0.22) | 1112 (0.21) | 7853 (0.24) | 8528 (0.25) |
| | | Q3 | 9606 (0.29) | 10295 (0.30) | 2273 (0.33) | 1660 (0.32) | 9903 (0.30) | 10451 (0.30) |
| | | Q4 | 10293 (0.31) | 10801 (0.31) | 2510 (0.36) | 1956 (0.38) | 10826 (0.33) | 11721 (0.34) |
| | | Missing | | | | | | |
| | Externalizing problems | SDQ externalizing score not in top 10th percentile | 15087 (0.46) | 15526 (0.45) | 6155 (0.88) | 4742 (0.92) | 28404 (0.87) | 31433 (0.90) |
| | | SDQ externalizing score in top 10th percentile | 2257 (0.07) | 1772 (0.05) | 801 (0.11) | 416 (0.08) | 4408 (0.13) | 3348 (0.10) |
| | | Missing | 15468 (0.47) | 17483 (0.50) | 33 (0.00) | 23 (0.00) | | |
| | Internalizing problems | SDQ internalizing score not in top 10th percentile | 14962 (0.46) | 14974 (0.43) | 6030 (0.86) | 4509 (0.87) | 27825 (0.85) | 29869 (0.86) |
| | | SDQ internalizing score in top 10th percentile | 2382 (0.07) | 2324 (0.07) | 926 (0.13) | 649 (0.13) | 4987 (0.15) | 4912 (0.14) |
| | | Missing | 15468 (0.47) | 17483 (0.50) | 33 (0.00) | 23 (0.00) | | |
| | Prosocial problems | SDQ prosocial score not in bottom 10th percentile | 14835 (0.45) | 14975 (0.43) | 5927 (0.85) | 4491 (0.87) | 27748 (0.85) | 30187 (0.87) |
| | | SDQ prosocial score in bottom 10th percentile | 2509 (0.08) | 2323 (0.07) | 1029 (0.15) | 667 (0.13) | 5064 (0.15) | 4594 (0.13) |
| | | Missing | 15468 (0.47) | 17483 (0.50) | 33 (0.00) | 23 (0.00) | | |
| | Physical activity | Inactive | 2568 (0.08) | 1275 (0.04) | 996 (0.14) | 437 (0.08) | 4850 (0.15) | 3071 (0.09) |

| Tier | Variable | Level | Female study population | Male study population | Female complete participants | Male complete participants | Female analysis sample | Male analysis sample |
|---|---|---|---|---|---|---|---|---|
| | | Lightly active | 8008 (0.24) | 7222 (0.21) | 3083 (0.44) | 2264 (0.44) | 15357 (0.47) | 16255 (0.47) |
| | | Moderately active | 6494 (0.20) | 6976 (0.20) | 2468 (0.35) | 2030 (0.39) | 11389 (0.35) | 13979 (0.40) |
| | | Vigorously active | 733 (0.02) | 858 (0.02) | 256 (0.04) | 232 (0.04) | 1216 (0.04) | 1476 (0.04) |
| | | Missing | 15009 (0.46) | 18450 (0.53) | 186 (0.03) | 218 (0.04) | | |
| | Affective disorders | No diagnosis | 32446 (0.99) | 34327 (0.99) | 6927 (0.99) | 5126 (0.99) | 32440 (0.99) | 34354 (0.99) |
| | | One or more diagnoses | 366 (0.01) | 454 (0.01) | 62 (0.01) | 55 (0.01) | 372 (0.01) | 427 (0.01) |
| | Neurodevelopmental disorders | No diagnosis | 32312 (0.98) | 33108 (0.95) | 6925 (0.99) | 5031 (0.97) | 32378 (0.99) | 33421 (0.96) |
| | | One or more diagnoses | 500 (0.02) | 1673 (0.05) | 64 (0.01) | 150 (0.03) | 434 (0.01) | 1360 (0.04) |
| | Other psychiatric diagnoses | No diagnosis | 32631 (0.99) | 34386 (0.99) | 6966 (1.00) | 5149 (0.99) | 32659 (1.00) | 34503 (0.99) |
| | | One or more diagnoses | 181 (0.01) | 395 (0.01) | 23 (0.00) | 32 (0.01) | 153 (0.00) | 278 (0.01) |
| | Screen time | 2 or less hours | 7108 (0.22) | 2857 (0.08) | 2723 (0.39) | 852 (0.16) | 12734 (0.39) | 5712 (0.16) |
| | | 2 to 4 hours | 7323 (0.22) | 6471 (0.19) | 2764 (0.40) | 1941 (0.37) | 13106 (0.40) | 13424 (0.39) |
| | | 4 to 6 hours | 2512 (0.08) | 3737 (0.11) | 922 (0.13) | 1148 (0.22) | 4575 (0.14) | 7841 (0.23) |
| | | 6 or more hours | 1356 (0.04) | 3800 (0.11) | 474 (0.07) | 1101 (0.21) | 2397 (0.07) | 7804 (0.22) |
| | | Missing | 14513 (0.44) | 17916 (0.52) | 106 (0.02) | 139 (0.03) | | |
| | Sleep time | 19:00 or earlier | 44 (0.00) | 46 (0.00) | 15 (0.00) | 10 (0.00) | 81 (0.00) | 64 (0.00) |
| | | 19:30 to 20:00 | 873 (0.03) | 938 (0.03) | 375 (0.05) | 311 (0.06) | 1761 (0.05) | 2063 (0.06) |
| | | 20:30 to 21:00 | 9214 (0.28) | 8541 (0.25) | 3611 (0.52) | 2620 (0.51) | 16794 (0.51) | 17849 (0.51) |
| | | 21:30 to 22:00 | 7439 (0.23) | 6806 (0.20) | 2652 (0.38) | 1946 (0.38) | 12764 (0.39) | 13375 (0.38) |
| | | 22:30 to 23:00 | 831 (0.03) | 732 (0.02) | 236 (0.03) | 182 (0.04) | 1259 (0.04) | 1324 (0.04) |
| | | 23:30 or later | 77 (0.00) | 74 (0.00) | 23 (0.00) | 12 (0.00) | 153 (0.00) | 106 (0.00) |

| Tier | Variable | Level | Female study population | Male study population | Female complete participants | Male complete participants | Female analysis sample | Male analysis sample |
|---|---|---|---|---|---|---|---|---|
| | | Missing | 14334 (0.44) | 17644 (0.51) | 77 (0.01) | 100 (0.02) | | |
| **Late adolescence** | Weight | BMI below 18.5 | 1918 (0.06) | 1231 (0.04) | 688 (0.10) | 469 (0.09) | 3229 (0.10) | 3080 (0.09) |
| | | BMI 18.5 - 24.9 | 13681 (0.42) | 9929 (0.29) | 4983 (0.71) | 3564 (0.69) | 24559 (0.75) | 26387 (0.76) |
| | | BMI 25.0 - 29.9 | 2421 (0.07) | 1785 (0.05) | 722 (0.10) | 548 (0.11) | 3765 (0.11) | 4157 (0.12) |
| | | BMI above 30.0 | 863 (0.03) | 535 (0.02) | 205 (0.03) | 129 (0.02) | 1259 (0.04) | 1157 (0.03) |
| | | Missing | 13929 (0.42) | 21301 (0.61) | 391 (0.06) | 471 (0.09) | | |
| | Major Depression Inventory score | No depressive symptoms | 15222 (0.46) | 13095 (0.38) | 5392 (0.77) | 4543 (0.88) | 25290 (0.77) | 31847 (0.92) |
| | | Mild depression | 1780 (0.05) | 597 (0.02) | 565 (0.08) | 182 (0.04) | 2842 (0.09) | 1299 (0.04) |
| | | Moderate depression | 1242 (0.04) | 372 (0.01) | 400 (0.06) | 109 (0.02) | 2028 (0.06) | 792 (0.02) |
| | | Severe depression | 1611 (0.05) | 382 (0.01) | 459 (0.07) | 110 (0.02) | 2652 (0.08) | 843 (0.02) |
| | | Missing | 12957 (0.39) | 20335 (0.58) | 173 (0.02) | 237 (0.05) | | |
| | Mental well-being (SWEMBS) | Probable anxiety or depression | 1045 (0.03) | 457 (0.01) | 302 (0.04) | 130 (0.03) | 1697 (0.05) | 946 (0.03) |
| | | Medium possible anxiety or depression | 2417 (0.07) | 1141 (0.03) | 735 (0.11) | 345 (0.07) | 3792 (0.12) | 2592 (0.07) |
| | | Good mental well-being | 17254 (0.53) | 13947 (0.40) | 5952 (0.85) | 4706 (0.91) | 27323 (0.83) | 31243 (0.90) |
| | | Missing | 12096 (0.37) | 19236 (0.55) | | | | |
| | Adversity: family | No occurrence | 23526 (0.72) | 25215 (0.72) | 5459 (0.78) | 4106 (0.79) | 24354 (0.74) | 26195 (0.75) |
| | | Minimum one occurrence | 9286 (0.28) | 9566 (0.28) | 1530 (0.22) | 1075 (0.21) | 8458 (0.26) | 8586 (0.25) |
| | Adversity: loss | No occurrence | 28779 (0.88) | 30456 (0.88) | 6194 (0.89) | 4606 (0.89) | 28754 (0.88) | 30541 (0.88) |

| Tier | Variable | Level | Female study population | Male study population | Female complete participants | Male complete participants | Female analysis sample | Male analysis sample |
|---|---|---|---|---|---|---|---|---|
| | | Minimum one occurrence | 4033 (0.12) | 4325 (0.12) | 795 (0.11) | 575 (0.11) | 4058 (0.12) | 4240 (0.12) |
| | Adversity: material | No occurrence | 31041 (0.95) | 32934 (0.95) | 6705 (0.96) | 5000 (0.97) | 31073 (0.95) | 33240 (0.96) |
| | | Minimum one occurrence | 1771 (0.05) | 1847 (0.05) | 284 (0.04) | 181 (0.03) | 1739 (0.05) | 1541 (0.04) |
| | Externalizing problems | SDQ externalizing score not in top 10th percentile | 17300 (0.53) | 12042 (0.35) | 6091 (0.87) | 4240 (0.82) | 29343 (0.89) | 30527 (0.88) |
| | | SDQ externalizing score in top 10th percentile | 2121 (0.06) | 1918 (0.06) | 632 (0.09) | 585 (0.11) | 3469 (0.11) | 4254 (0.12) |
| | | Missing | 13391 (0.41) | 20821 (0.60) | 266 (0.04) | 356 (0.07) | | |
| | Internalizing problems | SDQ internalizing score not in top 10th percentile | 17059 (0.52) | 12454 (0.36) | 5972 (0.85) | 4386 (0.85) | 28594 (0.87) | 31313 (0.90) |
| | | SDQ internalizing score in top 10th percentile | 2446 (0.07) | 1599 (0.05) | 765 (0.11) | 460 (0.09) | 4218 (0.13) | 3468 (0.10) |
| | | Missing | 13307 (0.41) | 20728 (0.60) | 252 (0.04) | 335 (0.06) | | |
| | Prosocial problems | SDQ prosocial score not in bottom 10th percentile | 17294 (0.53) | 12370 (0.36) | 5977 (0.86) | 4279 (0.83) | 29130 (0.89) | 30851 (0.89) |
| | | SDQ prosocial score in bottom 10th percentile | 2211 (0.07) | 1683 (0.05) | 760 (0.11) | 567 (0.11) | 3682 (0.11) | 3930 (0.11) |
| | | Missing | 13307 (0.41) | 20728 (0.60) | 252 (0.04) | 335 (0.06) | | |
| | Life satisfaction (Cantril) | Low quality of life | 4481 (0.14) | 2253 (0.06) | 1364 (0.20) | 659 (0.13) | 6968 (0.21) | 4775 (0.14) |
| | | Medium quality of life | 13286 (0.40) | 9578 (0.28) | 4514 (0.65) | 3196 (0.62) | 20999 (0.64) | 21242 (0.61) |
| | | High quality of life | 3131 (0.10) | 3957 (0.11) | 1111 (0.16) | 1326 (0.26) | 4845 (0.15) | 8764 (0.25) |
| | | Missing | 11914 (0.36) | 18993 (0.55) | | | | |
| | Household income | Q1 | 5396 (0.16) | 4915 (0.14) | 740 (0.11) | 426 (0.08) | 4543 (0.14) | 3817 (0.11) |

| Tier | Variable | Level | Female study population | Male study population | Female complete participants | Male complete participants | Female analysis sample | Male analysis sample |
|---|---|---|---|---|---|---|---|---|
| | | Q2 | 7900 (0.24) | 8414 (0.24) | 1585 (0.23) | 1123 (0.22) | 8033 (0.24) | 8313 (0.24) |
| | | Q3 | 9291 (0.28) | 10346 (0.30) | 2226 (0.32) | 1643 (0.32) | 9630 (0.29) | 10704 (0.31) |
| | | Q4 | 10192 (0.31) | 11083 (0.32) | 2438 (0.35) | 1989 (0.38) | 10606 (0.32) | 11947 (0.34) |
| | | Missing | 33 (0.00) | 23 (0.00) | | | | |
| | Affective disorders | No diagnosis | 29911 (0.91) | 33352 (0.96) | 6492 (0.93) | 5014 (0.97) | 29996 (0.91) | 33467 (0.96) |
| | | One or more diagnoses | 2901 (0.09) | 1429 (0.04) | 497 (0.07) | 167 (0.03) | 2816 (0.09) | 1314 (0.04) |
| | Neurodevelopmental | No diagnosis | 31268 (0.95) | 32560 (0.94) | 6748 (0.97) | 4989 (0.96) | 31304 (0.95) | 33150 (0.95) |
| | | One or more diagnoses | 1544 (0.05) | 2221 (0.06) | 241 (0.03) | 192 (0.04) | 1508 (0.05) | 1631 (0.05) |
| | Other psychiatric diagnoses | No diagnosis | 31646 (0.96) | 34361 (0.99) | 6808 (0.97) | 5150 (0.99) | 31754 (0.97) | 34506 (0.99) |
| | | One or more diagnoses | 1166 (0.04) | 420 (0.01) | 181 (0.03) | 31 (0.01) | 1058 (0.03) | 275 (0.01) |
| | Psychotic disorders | No diagnosis | 32532 (0.99) | 34618 (1.00) | 6949 (0.99) | 5160 (1.00) | 32570 (0.99) | 34598 (0.99) |
| | | One or more diagnoses | 280 (0.01) | 163 (0.00) | 40 (0.01) | 21 (0.00) | 242 (0.01) | 183 (0.01) |
| | Sleep time | 20:00 or earlier | 68 (0.00) | 62 (0.00) | 19 (0.00) | 25 (0.00) | 103 (0.00) | 176 (0.01) |
| | | 21:00 to 22:00 | 7780 (0.24) | 3773 (0.11) | 2782 (0.40) | 1356 (0.26) | 12909 (0.39) | 9046 (0.26) |
| | | 23:00 to 00:00 | 11366 (0.35) | 9475 (0.27) | 3803 (0.54) | 3198 (0.62) | 18232 (0.56) | 22193 (0.64) |
| | | 01:00 to 02:00 | 969 (0.03) | 1457 (0.04) | 272 (0.04) | 439 (0.08) | 1455 (0.04) | 3195 (0.09) |
| | | 03:00 or later | 85 (0.00) | 134 (0.00) | 19 (0.00) | 23 (0.00) | 113 (0.00) | 171 (0.00) |
| | | Missing | 12544 (0.38) | 19880 (0.57) | 94 (0.01) | 140 (0.03) | | |

*Table S2: Distributions of all included variables and levels in the study population, the full-participant population, and the cloned analysis population (to account for selection), stratified by sex.* Table S1 *includes details on the categorization of the levels of all variables*

**DISCED refers to the Danish International Standard Classification of Education (Classification on Education (DISCED-15), which is Denmark's national adaptation of UNESCO's International Standard Classification of Education (ISCED). This variable is categorized as Low Education (DISCED 10, 20); Medium Education (DISCED 30, 40) High Education (DISCED 50, 60, 70, 80).*

*Abbreviations: BMI: Body Mass Index, SDQ: Strengths and Difficulties Questionnaire; DAWBA: Development and Well-Being Assessment; SiC: Stress in Children Questionnaire*

| Threshold | Boys (also in main) | Girls (also in main) |
|---|---|---|
| 1% | 1294 (300) | 1269 (298) |
| 5% | 891 (290) | 845 (296) |
| 10% | 675 (272) | 652 (282) |
| 25% | 401 (225) | 364 (238) |
| 50% | 189 (150) | 217 (180) |
| 75% | 106 (94) | 125 (118) |
| 90% | 75 (68) | 86 (86) |
| 95% | 58 (58) | 70 (70) |
| 99% | 43 (43) | 59 (59) |
| 100% | 39 (39) | 51 (51) |

*Table S3: Number of unique edges across the 100 bootstrap repetitions for both girls and boys, and how many of these are present in the TMPDAG from the main analysis. Each bootstrap repetition contains 300 edges (+/- 1). We report the number of unique edges for different levels of thresholds. Such that the first row counts the number of distinct edges that where present in at least 1% of the bootstrap repetitions and the last row counts the number of distinct edges that show up in every bootstrap repetition, for girls and boys separately. We count an undirected edge, a directed edge and an edge directed opposite as three unique edges.*

| Sex | ***Threshold for bootstrap repetitions*** | Weight process | Mental health process |
|---|---|---|---|
| **Girls** | **50%** | 38 | 56 |
| | **95%** | 18 | 18 |
| **Boys** | **50%** | 38 | 70 |
| | **95%** | 14 | 18 |

*Table S4: Number of pathways between all weight variables and all well-being variables.. Only count those where there exists a pathway in over 50% or 95% of the bootstrap repetitions. The reported numbers correspond to number of edges in Figures S3-S6.*

| Skipping pattern | Dataset | Pregnancy | At birth | Infancy | Childhood | Early adol. | Late adol. | $n_{boys}$ | $n_{girls}$ |
|---|---|---|---|---|---|---|---|---|---|
| Complete participants | $D_{complete}$ | | | | | | | 5181 | 6989 |
| Pattern 1 | $D_1$ | | | *Skipped* | *Skipped* | *Skipped* | *Skipped* | 3583 | 2451 |
| Pattern 2 | $D_2$ | | | *Skipped* | *Skipped* | *Skipped* | | 1510 | 2026 |
| Pattern 3 | $D_3$ | | | *Skipped* | *Skipped* | | *Skipped* | 582 | 397 |
| Pattern 4 | $D_4$ | | | *Skipped* | *Skipped* | | | 518 | 827 |
| Pattern 5 | $D_5$ | | | *Skipped* | | *Skipped* | *Skipped* | 1931 | 1216 |
| Pattern 6 | $D_6$ | | | *Skipped* | | *Skipped* | | 1265 | 1460 |
| Pattern 7 | $D_7$ | | | *Skipped* | | | *Skipped* | 1921 | 1172 |
| Pattern 8 | $D_8$ | | | *Skipped* | | | | 2159 | 2906 |
| Pattern 9 | $D_9$ | | | | *Skipped* | *Skipped* | *Skipped* | 3245 | 2180 |
| Pattern 10 | $D_{10}$ | | | | *Skipped* | *Skipped* | | 1936 | 2557 |
| Pattern 11 | $D_{11}$ | | | | *Skipped* | | *Skipped* | 856 | 553 |
| Pattern 12 | $D_{12}$ | | | | *Skipped* | | | 872 | 1407 |
| Pattern 13 | $D_{13}$ | | | | | *Skipped* | *Skipped* | 3111 | 1757 |
| Pattern 14 | $D_{14}$ | | | | | *Skipped* | | 2104 | 2544 |
| Pattern 15 | $D_{15}$ | | | | | | *Skipped* | 4007 | 2370 |

*Table S5: Data collection skipping patterns observed in the study population. Empty cells represent non-skipped data collections. "Adol." abbreviates "adolescence". $n_{boy}$s and $n_{girls}$ reports the number of individuals with each skipping pattern in the dataset for boys and girls, respectively.*

**A)**

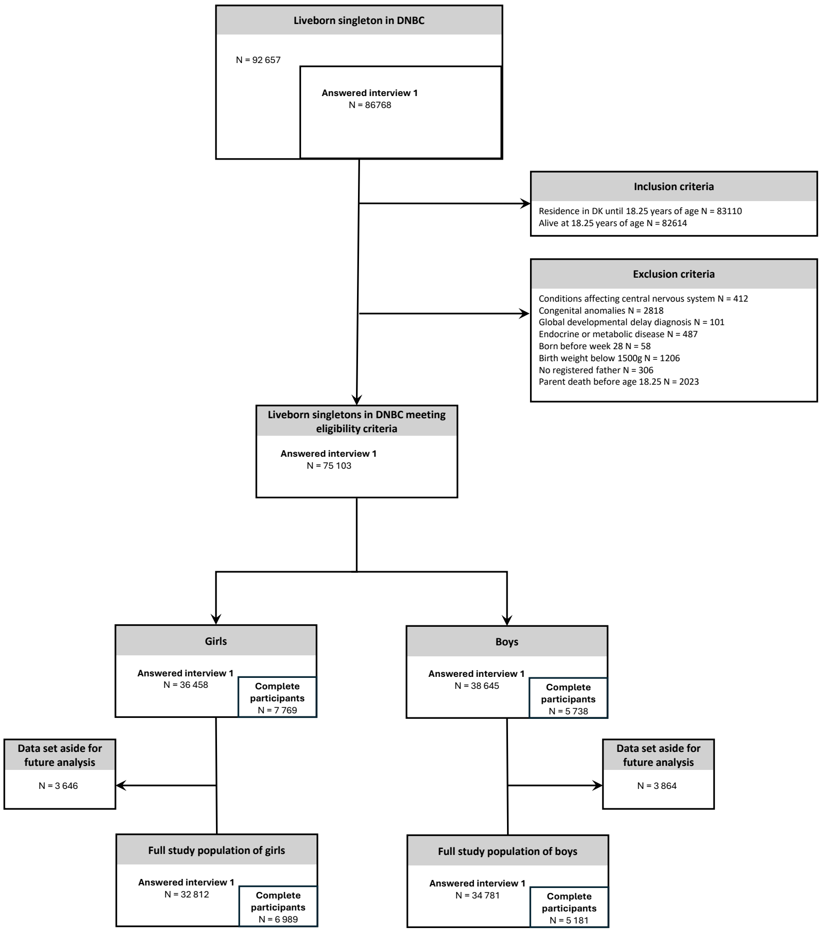


Figure S1: Data flowchart describing the number of individuals in the different populations, the inclusion and exclusion mechanisms and the random data split.

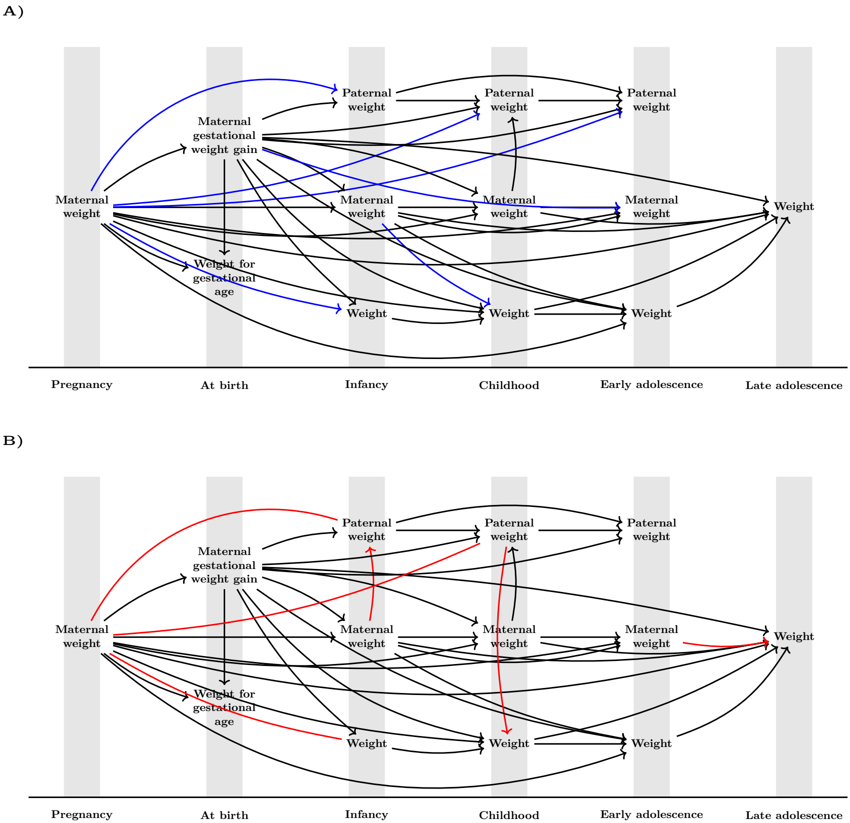


Figure S2: Weight development processes for boys (A) and girls (B) with edges for pathways present in over 50% of the bootstrap repetitions. Black edges are shared by boys and girls; red edges are unique to girls and blue are unique to boys. Directed edges represent directed causal paths that are not exclusively mediated via any variables shown in the graph. Undirected edges represent undirected or partially directed causal paths that are not exclusively mediated via any variables shown in the graph.

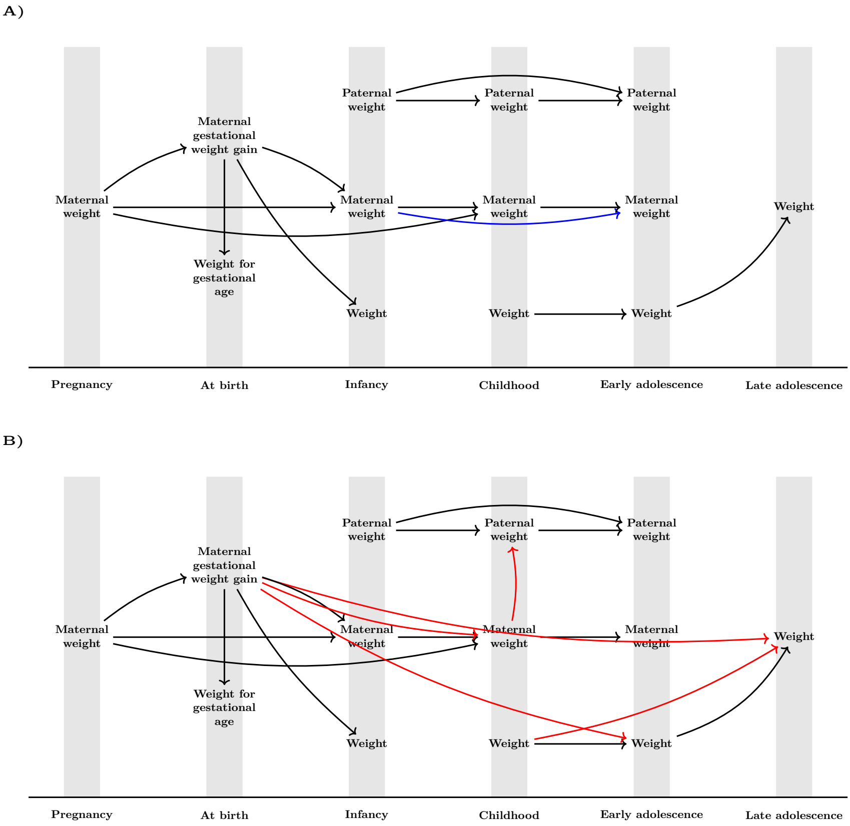


Figure S3: Weight development processes for boys (A) and girls (B) with edges for pathways present in minimum 95% of the bootstrap repetitions. Black edges are shared by boys and girls; red edges are unique to girls and blue are unique to boys. Directed edges represent directed causal paths that are not exclusively mediated via any variables shown in the graph. Undirected edges represent undirected or partially directed causal paths that are not exclusively mediated via any variables shown in the graph.

**A)**

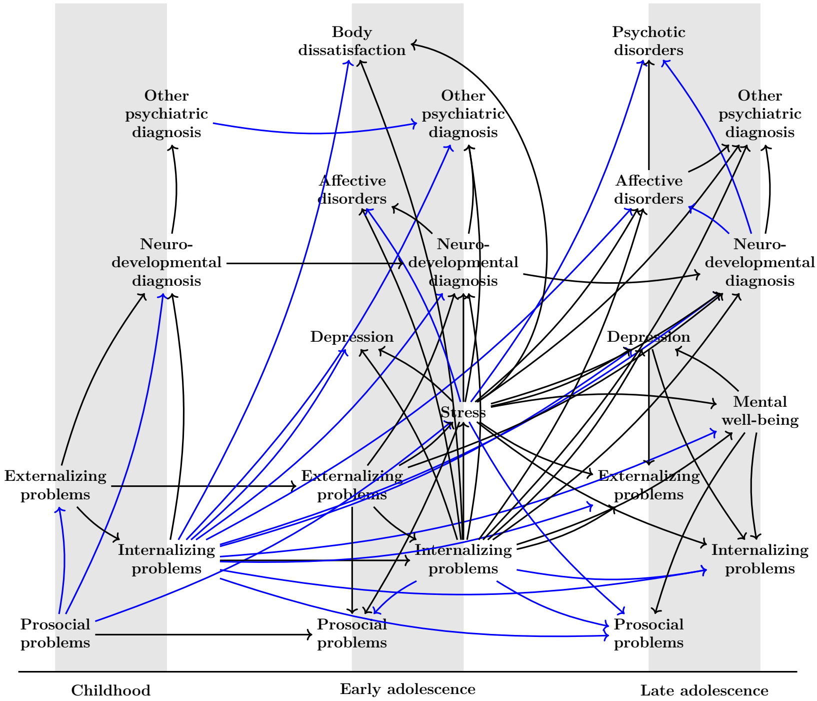


**B)**

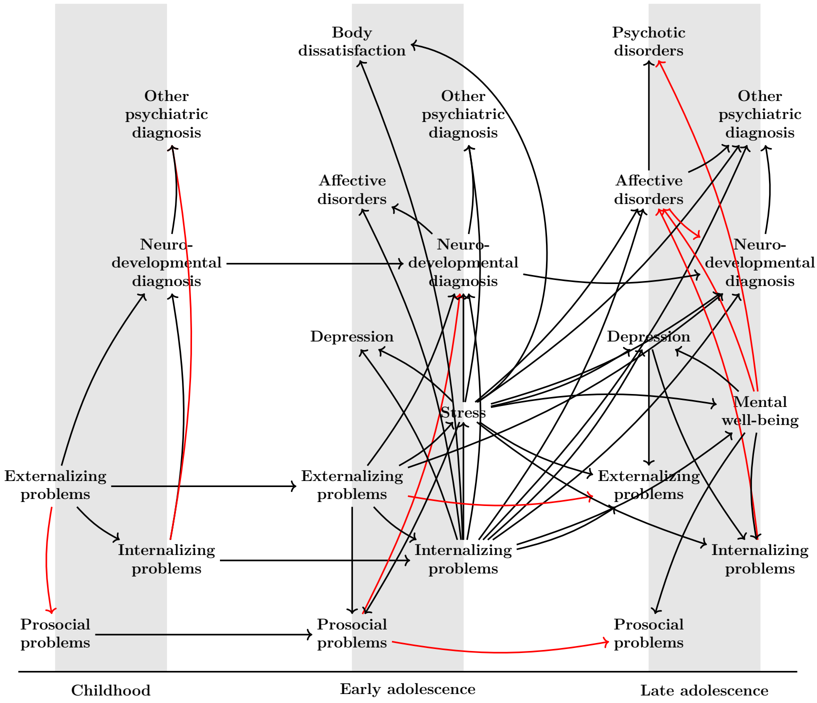


Figure S4: Mental health development processes for boys (A) and girls (B) with edges for pathways present in over 50% of the bootstrap repetitions. Black edges are shared by boys and girls; red edges are unique to girls and blue are unique to boys. Directed edges represent directed causal paths that are not exclusively mediated via any variables shown in the graph.

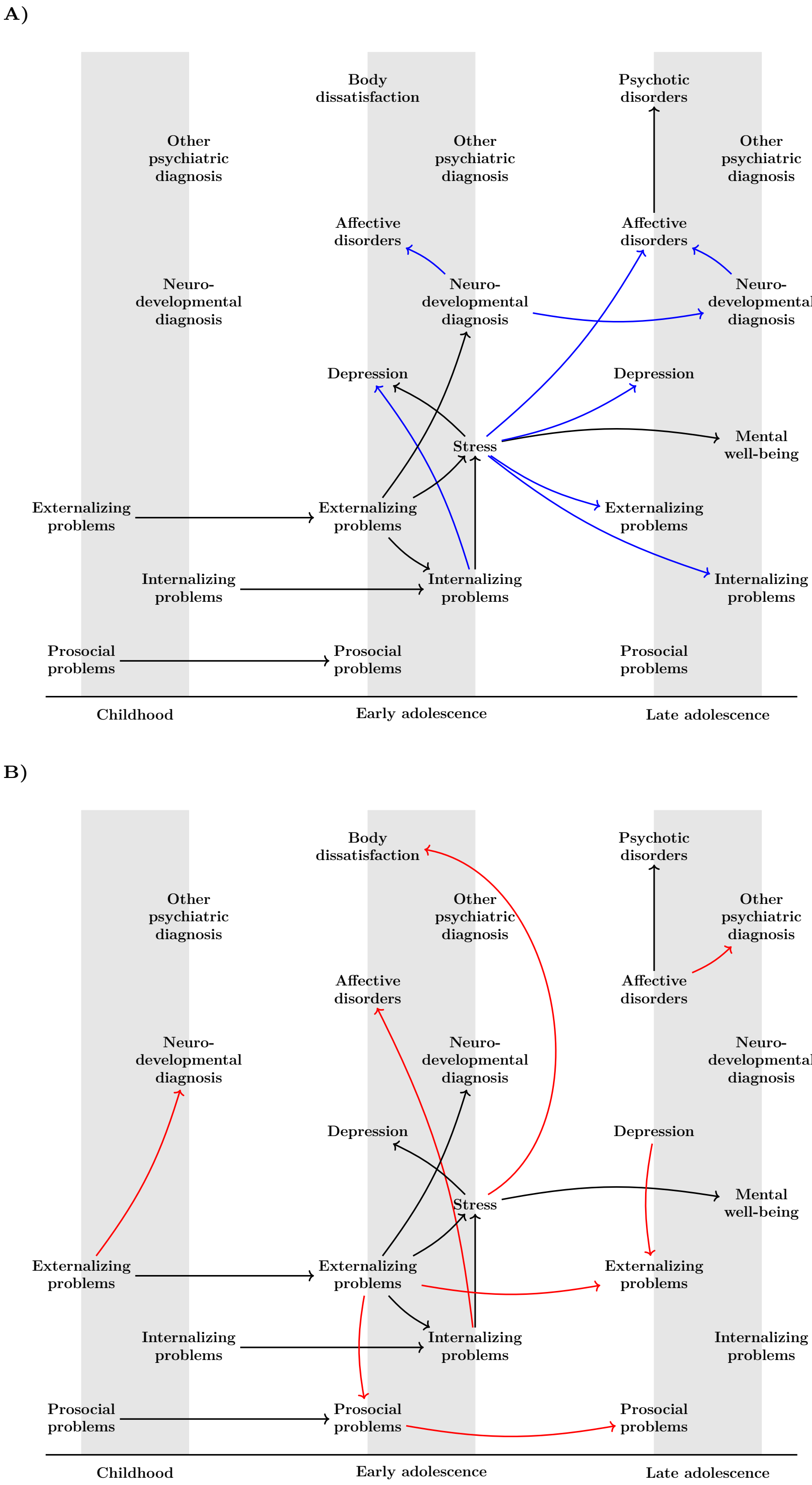


Figure S5: Mental health development processes for boys (A) and girls (B) with edges for pathways present in minimum 95% of the bootstrap repetitions. Black edges are shared by boys and girls; red edges are unique to girls and blue are unique to boys. Directed edges represent directed causal paths that are not exclusively mediated via any variables shown in the graph.

**A)**

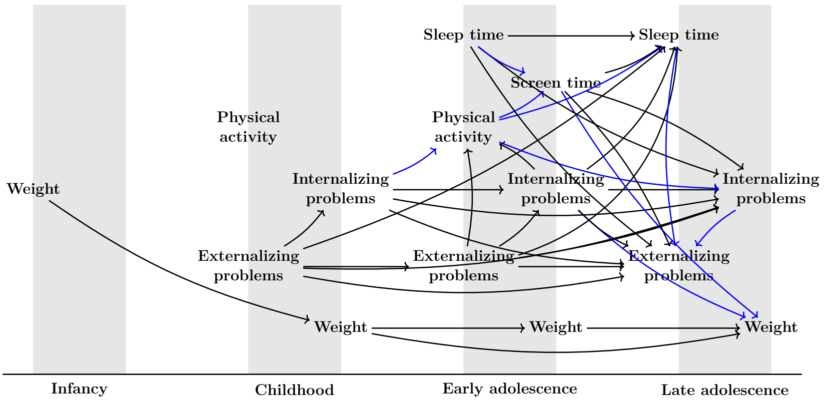


**B)**

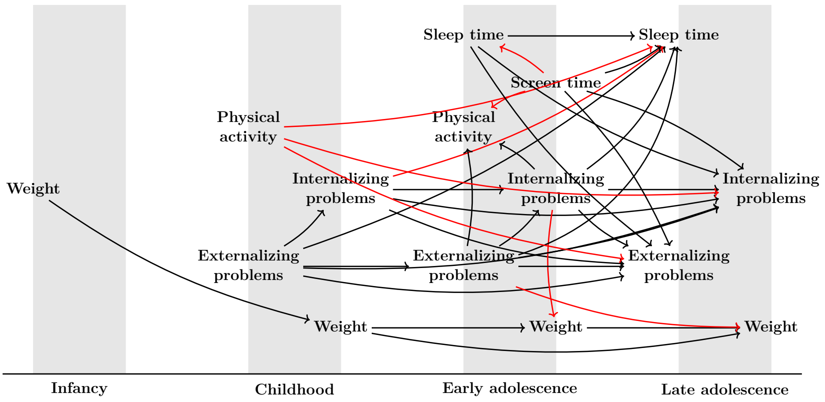


Figure S6: Interplay between weight development, internalizing/externalizing problems and selected life-style risk factors for boys (A) and girls (B) with edges for pathways present in over 50% of the bootstrap repetitions. Black edges are shared by boys and girls; red edges are unique to girls and blue are unique to boys. Directed edges represent directed causal paths that are not exclusively mediated via any variables shown in the graph. No undirected edges were found. We omit variables from "At birth as there were no paths between these and any other node in the graph.

**A)**

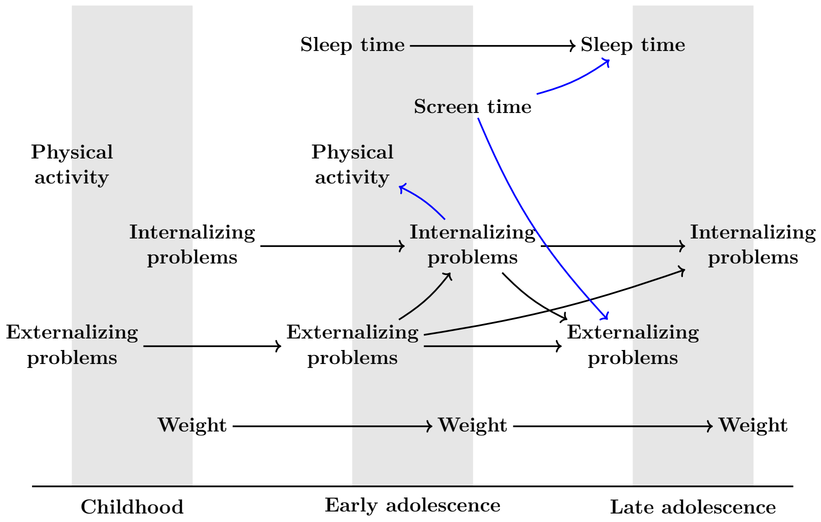


**B)**

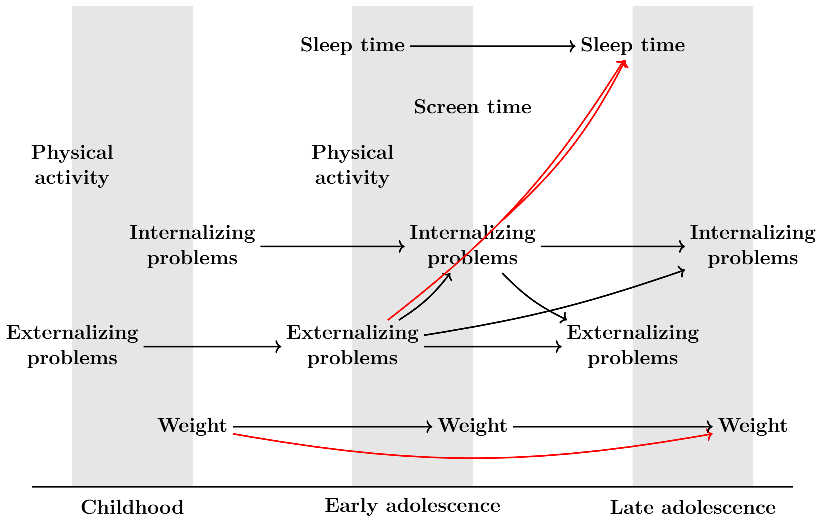


Figure S7: Interplay between weight development, internalizing/externalizing problems and selected life-style risk factors for boys (A) and girls (B) with edges for pathways present in minimum 95% of the bootstrap repetitions. Black edges are shared by boys and girls; red edges are unique to girls and blue are unique to boys. Directed edges represent directed causal paths that are not exclusively mediated via any variables shown in the graph. No undirected edges were found. We omit variables from “At birth” and “Infancy” as there were no paths between these and any other node in the graph.

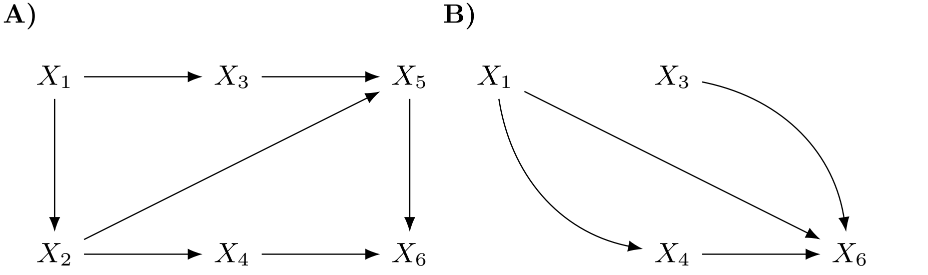


Figure S8: Subgraph example. A TMPDAG (A) and its corresponding subgraph (B) over variables $S = (X_1, X_3, X_4, X_6)$.
